# Distinct Stem Cell Identities Converge into Shared Erythroid Stress in ERCC6L2 Disease and Shwachman-Diamond Syndrome

**DOI:** 10.1101/2025.09.11.25335313

**Authors:** Laura Langohr, Ilse Kaaja, Suvi Douglas, Hanna Nebelung, Jessica Koski, Ina Ikonen, Lotta Katainen, Katri Maljanen, Marja Hakkarainen, Tuulia Räisänen, Riitta Niinimäki, Sakari Kakko, Timo Siitonen, Sadiksha Adhikari, Markus Vähä-Koskela, Caroline A. Heckman, Jenni Lahtela, Ulla Wartiovaara-Kautto, Esa Pitkänen, Outi Kilpivaara

## Abstract

ERCC6L2 disease (ED) is a rare bone marrow failure syndrome caused by biallelic germline mutations in *ERCC6L2*. ED leads to accumulation of somatic *TP53* mutations, myelodysplastic syndrome, and acute myeloid leukemia (AML) with erythroid predominance and poor prognosis. Given the global challenge of treating *TP53*-mutated AML, ED provides a unique opportunity to study early events leading to high-risk leukemia. While ERCC6L2 is implicated in DNA replication and repair, the transcriptomic events underlying delayed erythropoiesis and leukemic progression remain largely undefined. To delineate these processes, we leverage bulk and single-cell transcriptomics of patient fibroblasts, bone marrow, and peripheral blood across disease stages, including single-cell *TP53* genotyping.

We identify disease-associated erythroid delay and ferroptotic stress emerging prior to *TP53* mutation, highlighting an early vulnerability in ED leukemogenesis. We compare ED to Shwachman-Diamond syndrome (SDS) to reveal shared and disease-specific transcriptional programs. *TP53* mutations in ED and SDS arise in hematopoietic stem and progenitor cells but do not independently drive changes in cell cycle or stress pathways during erythropoiesis. Both diseases converge in late erythropoiesis into a stress state characterized by ferroptotic signaling, G1 arrest, and *BCL2L1* upregulation. As a disease-specific pattern, ED shows aberrant erythroid priming and a differentiation block in *TP53*-mutated cells during leukemic transformation.

Taken together, we provide the first single-cell analyses in ED, define stress responses shared with SDS, and aberrant erythroid priming with *TP53*-driven differentiation arrest shaping progression toward erythroid leukemia. Our results give biological insights to guide therapy development aiming at intercepting disease evolution to treatment-refractory AML.

**Key points:**

1. ERCC6L2 disease patient cells engage maladaptive erythroid programs early in differentiation
2. Cell cycle and ferroptotic stress signatures persist despite somatic *TP53* mutations

## Introduction

ERCC6L2 disease (ED) is an inherited bone marrow failure (BMF) syndrome caused by biallelic germline mutations in *ERCC6L2*^1–3^. ED patients are predisposed to primarily erythroid, lineage-restricted myeloid malignancy^4,5^. ERCC6L2 functions in DNA repair^1,2,6–8^, centromere stability^9^, mitochondrial homeostasis^1^, regulation and facilitation of transcription^8^, and class-switch recombination^10^. Clinically, ED often presents with BMF characterized by mild to moderate peripheral blood cytopenias^5^. As the condition progresses, multiple *TP53*-mutated hematopoietic clones develop as somatic compensation to declining bone marrow (BM) function^11,12^. This evokes a high risk of loss of heterozygosity, and biallelic *TP53* inactivation, a pivotal event that propels the development of an aggressive, erythroid-predominant myeloid malignancy. Despite significant efforts to find cures for *TP53*-mutated myeloid malignancies, their prognosis is dismal^13^. Particularly for ED-related hematological malignancy if diagnosed at the final, leukemic stage, the outcomes are universally unfavorable^5^.

In light of these challenges, our study shifts focus from malignancy to earlier disease stages by investigating the transcriptomic landscape of *ERCC6L2*-deficient hematopoiesis, particularly during BMF. By identifying biological aberrations caused by *ERCC6L2* germline defects, we uncover insights for future studies on potential therapeutic targets. A recent study showed that *ERCC6L2* deficiency in hematopoietic stem and progenitor cells leads to delayed erythroid differentiation^14^. However, the contribution of somatic *TP53* mutations to this differentiation delay remains unclear. Our goal is to identify disease-specific transcriptional programs in ED by comparing it to Shwachman-Diamond syndrome (SDS), another inherited BMF syndrome with a propensity for *TP53*-mutated myeloid malignancies^11,15^. Leveraging an exceptionally comprehensive dataset for a rare disease, we analyzed erythropoiesis in ED patients by comparing their peripheral blood (PB) cell and BM cell transcriptomes to those of SDS patients and healthy controls. Within the BM, we further compared *TP53*-mutated and wild-type cells. We complemented these analyses with transcriptomic profiling of patient-derived fibroblasts to uncover germline-driven effects of *ERCC6L2* and *SBDS* deficiency, independent of somatic *TP53* alterations.

## Methods

Patient information, including *TP53* mutation status, was collected from the Finnish Hematological Registry and Biobank, and clinical repositories. The study was conducted in accordance with the Declaration of Helsinki. The study has been approved by Helsinki University Central Hospital ethics review committee (#206/13/03/03/2016, amendment 2023, and HRUHLAB2). All samples from living individuals are derived after written informed consent.

Patient samples were collected in conjunction with diagnostic samples. We examined the transcriptome of 16 BM samples from ten ED patients with homozygous Finnish founder mutation in *ERCC6L2* (NM_020207.7: c.1424delT, p.Ile475ThrfsTer36, rs768081343)^3–5,14^, six samples from five SDS patients, and 20 samples from 20 AML patients (none with erythroleukemia), together with 63 BM samples from eight controls^16–18^ and one erythroleukemia (AML M6) case^19^. We further examined the blood transcriptome of 28 samples from 12 ED patients, seven samples from five SDS patients, and 11 samples from nine controls; and the skin fibroblast transcriptome of 74 samples from eight ED patients, including one patient with a compound heterozygous mutation (the above-mentioned founder mutation and *ERCC6L2* NM_020207.7: c.4454_4455delGA, p.Arg1474ThrfsTer10, rs1553528966), 55 samples from four SDS patients, and 68 samples from eight controls (Figure 1A-B). ED and SDS patients carried up to four *TP53* mutations in their BM (Supplemental Table 1-2), most of them located in hotspots (Supplemental Figure 1).

**Figure 1.**
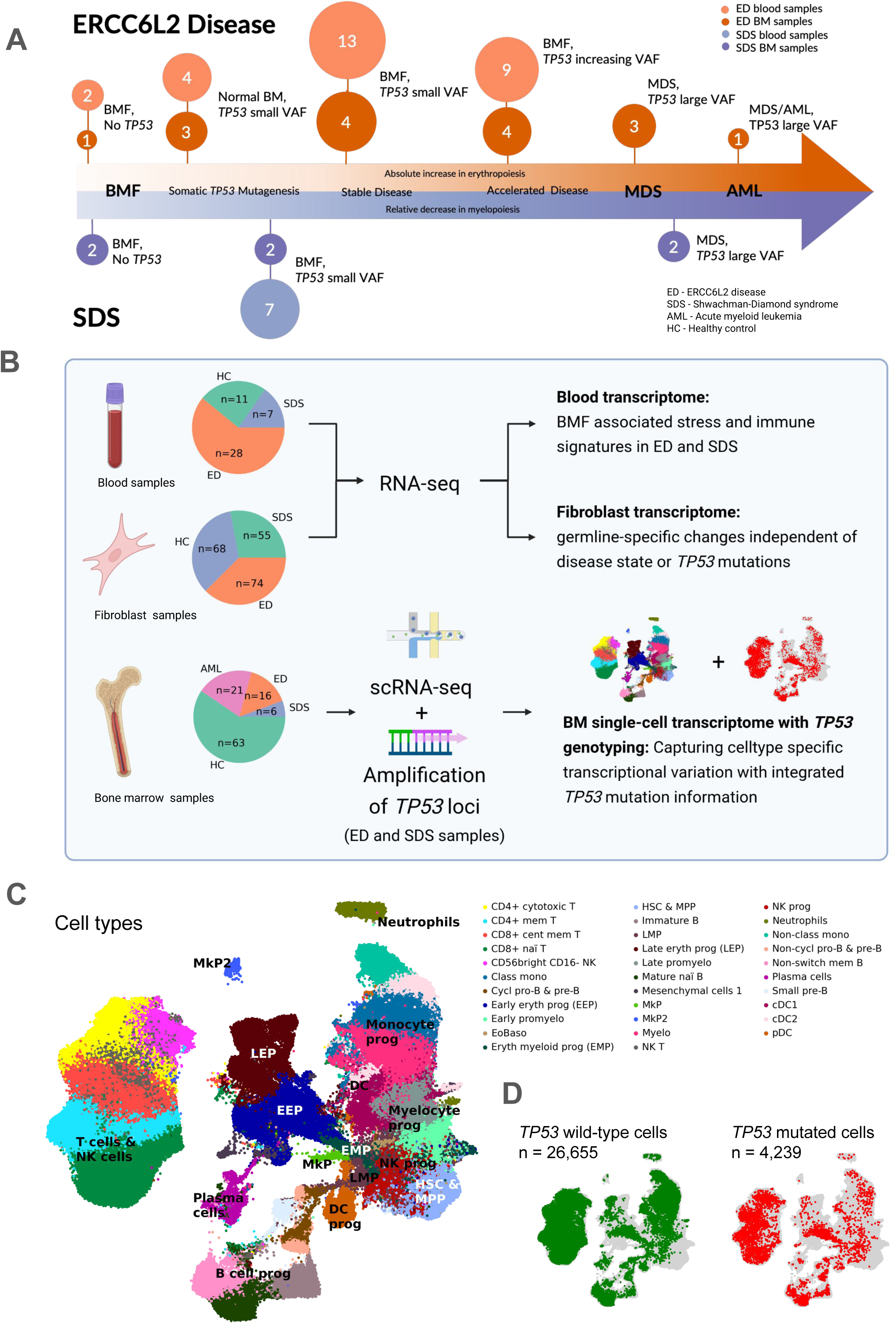
Data summary. **(A)** ED and SDS blood and BM samples included in this study, depicted at their different stages of the disease. Numbers denote the number of samples obtained from 10 ED and 5 SDS patients for BM, and from 12 ED and 5 SDS patients for blood. Colors depict the type of samples (bone marrow or blood). Samples with “no *TP53*” denote samples without somatic *TP53* mutations, and other samples depict cases with 1-4 *TP53* mutations. **(B)** Schematic of sample and data processing. Numbers denote the number of samples, amplification of *TP53* loci enabled targeted genotyping of known *TP53* mutation sites (Methods, Supplemental data). Image created with BioRender. **(C)** Detected cell types of our integrated single-cell transcriptomics data. **(D)** *TP53* mutation status of cells. Numbers denote the number of cells which were identified as *TP53*-mutated or *TP53* wild-type.

For BM samples, we applied single-cell RNA sequencing (scRNA-seq, 3’ 10X Genomics) and integrated the data using single-cell variational inference (scVI)^20^. Cell types were annotated with single-cell ANnotation using Variational Inference (scANVI)^21^ guided by a hematopoietic marker database^22^ (Supplemental Data, Supplemental Figure 2). This yielded 32 distinct cell types, including all major erythroid stages: hematopoietic stem cells and multipotent progenitor cells (HSC & MPP), erythroid-myeloid progenitors (EMP), early erythroid progenitors (EEP) and late erythroid progenitors (LEP) (Figure 1C).

To identify the *TP53*-mutated and wild-type cells, we applied targeted single-cell genotyping (scAmp-seq) within the same ED and SDS samples used for scRNA-seq, following established protocols^23,24^ (Supplemental Data, Supplemental Figure 3). *TP53* mutation statuses of cells were mapped to the scRNA-seq data via shared cell barcodes. We then performed the following analyses: differential expression (DE) with MAST^25^, pathway analyses using enrichR^26^ with Reactome database^27^, pseudotime using Genes2Genes^28^ with diffusion pseudotime^29^, gene regulatory network (GRN) using CellOracle^30^, copy number variation (CNV) with inferCNV^31^, and statistical tests.

For PB cells and fibroblasts, 3’ mRNA sequencing was performed with Illumina NextSeq 500/550 and aligned to GRCh38.p13^32^ (Supplemental data). DESeq2 (v.1.46.0)^33^ was used for DE (Supplemental Figure 4), and enrichR (v.3.2)^26^ for pathway analysis using Reactome 2024 database^27^. Across BM, PB, and fibroblast samples, genes were considered differentially expressed if *p_adj_*<0.05 and absolute log-fold change |log_2_FC|>0.25. A supervised gradient boosting model^34^ found no transcriptional markers of disease acceleration in ED PB or BM erythropoietic cells (Supplemental Figure 5). Detailed patient information is in Supplemental Table 1–3. Full methods are described in the Supplemental data.

### Data Sharing Statement

Count tables and processed scRNA-seq data from this study will be deposited at a repository. The raw data (FASTQ files) are not publicly available due to the ultra-rare nature of the disease, and to comply with the ethics approval and confidentiality agreements. Data may be obtained from the authors upon reasonable request when compatible with European General Data Protection Regulation (GDPR) and with permission from the University of Helsinki. Python and R scripts used to analyze and plot the data are available at GitHub: https://github.com/lalangohr/ERCC6L2-scRNA.

## Results

### Truncating *ERCC6L2* mutations escape nonsense mediated decay and exhibit expression beyond hematopoietic tissue

*ERCC6L2* p.Ile475ThrfsTer36 causes a premature termination codon (PTC) 105 nucleotides downstream of the mutation, with 3,122 nucleotides between the premature and normal stop codon. Classically, RNAs containing truncating mutations are targeted for degradation by nonsense mediated decay (NMD), but they may also escape NMD or be targeted with reduced NMD efficiency^35,36^. We examined the expression of *ERCC6L2* in the BM and PB cells (affected tissues) and skin fibroblasts (clinically unaffected tissue^5^). Despite the presence of biallelic truncating mutations in *ERCC6L2*, the transcript levels were detectable in ED samples, (Supplemental Figure 6A-B). We observed higher *ERCC6L2* expression in the compound heterozygous than in homozygous carriers in fibroblasts (Supplemental Figure 6A). In SDS, *SBDS* underexpression was visible in fibroblasts and BM compared to controls (Supplemental Figure 6A). Thus, the germline mutations in both *ERCC6L2* and *SBDS* genes lower the amount of their mRNA transcripts, compatible with partial NMD or altered mRNA surveillance.

### Somatic *TP53* mutations in ED and SDS arise in HSCs or early progenitor cells

To study the transcriptomic changes while distinguishing between *TP53*-mutated and wild-type cells, we integrated BM scRNA-seq data from ED, SDS, and AML patients, as well as controls and determined *TP53* mutation status for ED and SDS cells (Methods). The resulting dataset consists of 385,100 cells spanning 32 distinct cell types representing all hematopoietic cell lineages (Figure 1C) consistent with recent single-cell studies on BM^37,38^. ED and SDS in the BMF phase closely resembled controls in cell type proportions (Supplemental Figure 7), in line with clinical observations before malignant transformation^5,39^. We genotyped the *TP53* mutation status of 1.5–61.4% ED and SDS cells per sample and mutation site, identifying 26,655 *TP53* wild-type and 4,239 *TP53*-mutated cells (Figure 1C, Supplemental Methods, Supplemental Figure 3B). *TP53*-mutated cells were found in erythroid, myeloid as well as lymphoid cells (Figure 1D), demonstrating that *TP53* mutations affect the entire hematopoietic differentiation hierarchy, thus arising in HSCs or early progenitor cells (Supplemental Figure 8-9).

### Shared stress phenotypes in ED and SDS arise from distinct germline drivers

Since ED develops into erythroid leukemia specifically, we focused on the BM erythroid lineage^4,5^. When comparing differentially expressed genes (DEGs) between the two diseases, the most pronounced transcriptional differences between ED and SDS emerged in early hematopoietic stages with the disparity progressively diminishing during erythropoiesis (Pearson correlation *r*=0.5, r=0.6, *r*=0.64 and *r*=0.9 for HSC and MPP, EMP, EEP and LEP, respectively, Figure 2A, Supplemental Tables 5-8). Pathway analyses along erythropoiesis revealed shared stress signatures (Figure 2B, Supplemental Tables 9-12). Concordantly, transcriptomic analyses of PB cells revealed striking similarities between ED and SDS (Figure 2C, Supplemental Table 13), and showed upregulation of genes involved in cellular stress responses and immune pathways (Figure 2D, Supplemental Tables 14), consistent with prior studies on inherited BMF syndromes^40^. Patient-derived fibroblasts, lacking overt disease phenotypes or *TP53* mutations, exhibited transcriptional divergence between ED and SDS (Figure 2E, Supplemental Table 15). ED fibroblasts showed pathway enrichment related to amino acid sensing via the GCN2 response and axon guidance (Figure 2F, Supplemental Table 16), possibly reflecting generalized adaptation to chronic metabolic or proteostatic stress. SDS fibroblasts demonstrated broader alterations in RNA and protein metabolism (Figure 2F, Supplemental Table 16), pointing to distinct germline-driven effects on cellular homeostasis. These findings show that ED and SDS share stress-related transcriptional features in late erythroid progenitors and PB cells, while fibroblasts and early hematopoietic progenitors reveal germline-specific differences independent of disease or *TP53* mutation.

**Figure 2.**
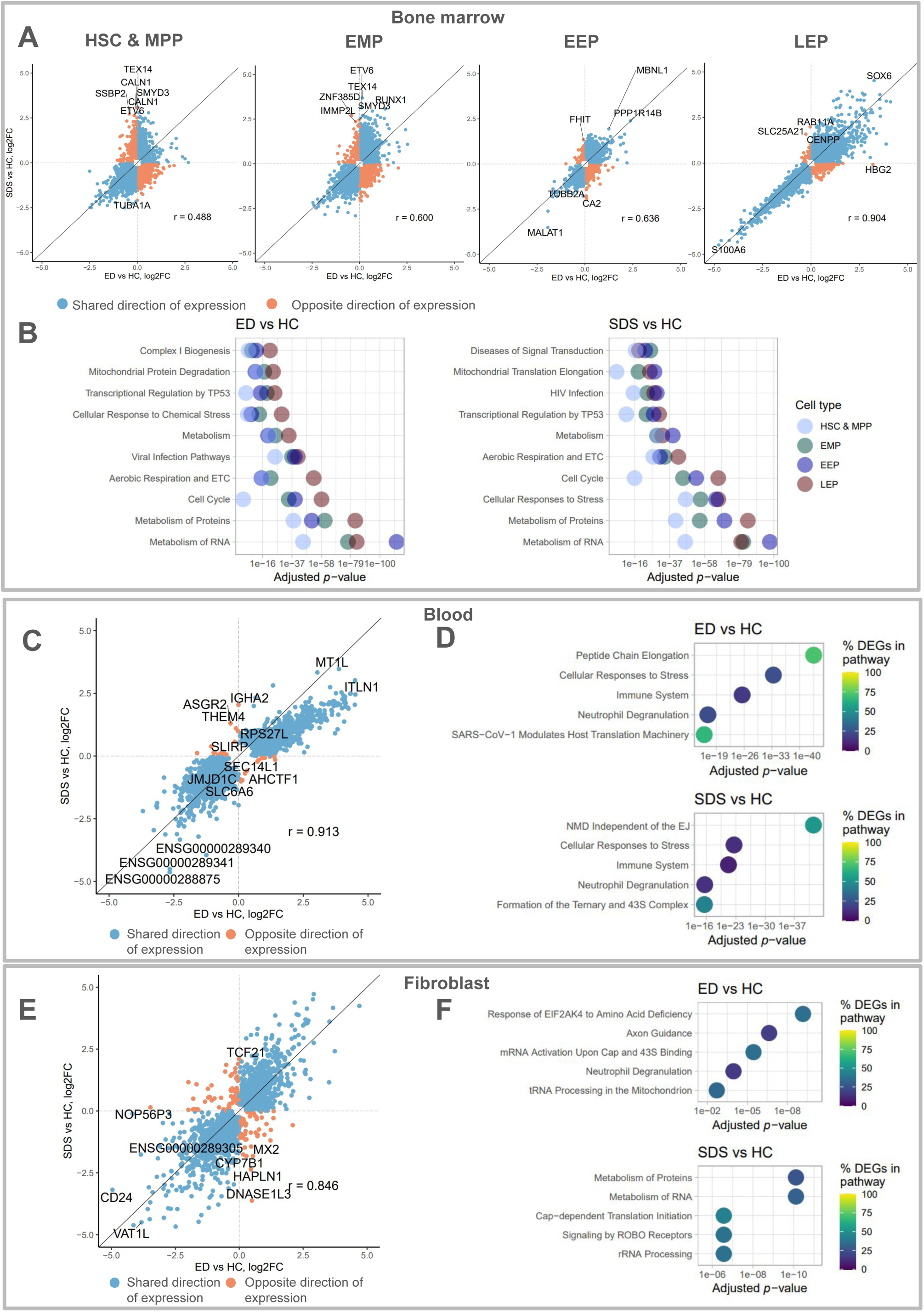
Transcriptional landscape of BM erythroid progenitors, peripheral blood cells and fibroblasts in ED compared to SDS. **(A)** Comparison of ED BMF and SDS BMF differentially expressed genes (DEGs) showing log_2_ fold changes (log2FC) of expression in ED BMF (n = 12 samples) and SDS BMF (n = 4) against healthy controls (n = 63) in bone marrow HSC and MPP, EMP, EEP, and LEP. **(B)** Top 10 non-redundant pathways across cell types for BM. Enriched pathways were sorted by FDR-adjusted p-values *p*_adj_. Redundant pathways (pathways containing DEGs of which more than half of the DEGs are members of a pathway with smaller *p*_adj_) and pathways not enriched for one of the cell types were filtered out. From the remaining pathways the top 10 based on the smallest *p*_adj_ across cell types were plotted. **(C)** Comparison of ED BMF and SDS BMF DEGs showing log_2_FC of expression in ED BMF (n = 28) and SDS BMF (n = 7) against healthy controls (n = 11) in blood samples. **(D)** Top five enriched pathways in ED BMF and SDS BMF compared to healthy controls in blood samples. **(E)** Comparison of ED and SDS DEGs showing log_2_FC of expression in ED (n = 74) and SDS (n = 55) against healthy controls (n = 68) in fibroblast samples. **(F)** Top five enriched pathways on ED and SDS compared to healthy controls in fibroblasts. r, Pearson correlation coefficient. DEGs, genes with *p*_adj_ < 0.05 in the DE analysis results. Enriched pathways, pathways with *p*_adj_ <0.05 in pathway analysis results.

### Aberrant Erythroid Priming Distinguishes ERCC6L2 Disease from SDS During Early Differentiation

To examine the transcriptional changes in ED and SDS in more detail, we first assessed the expression of erythropoiesis markers along erythropoietic differentiation (Figure 3A-C, Supplemental Methods). *CD34*, *GATA2* and *GATA1* followed their expected expression pattern in both diseases (Figure 3B-D), whereas *TFRC* and *SLC4A1* showed the most aberrant expression in ED BMF compared to controls (Figure 3B-D, expression dissimilarity *d*=1.03, 95% CI [0.97, 1.08] for *TFRC* and *d*=1.31, 95% CI [1.24, 1.38] for *SLC4A1,* Supplemental Table 17), which both showed aberrant expression also in SDS BMF (*d*=0.83, 95% CI [0.72, 0.93] for *TFRC* and *d*=1.04, 95% CI [0.93, 1.29] for *SLC4A1*, Supplemental Figure 10, Supplemental Table 17).

**Figure 3.**
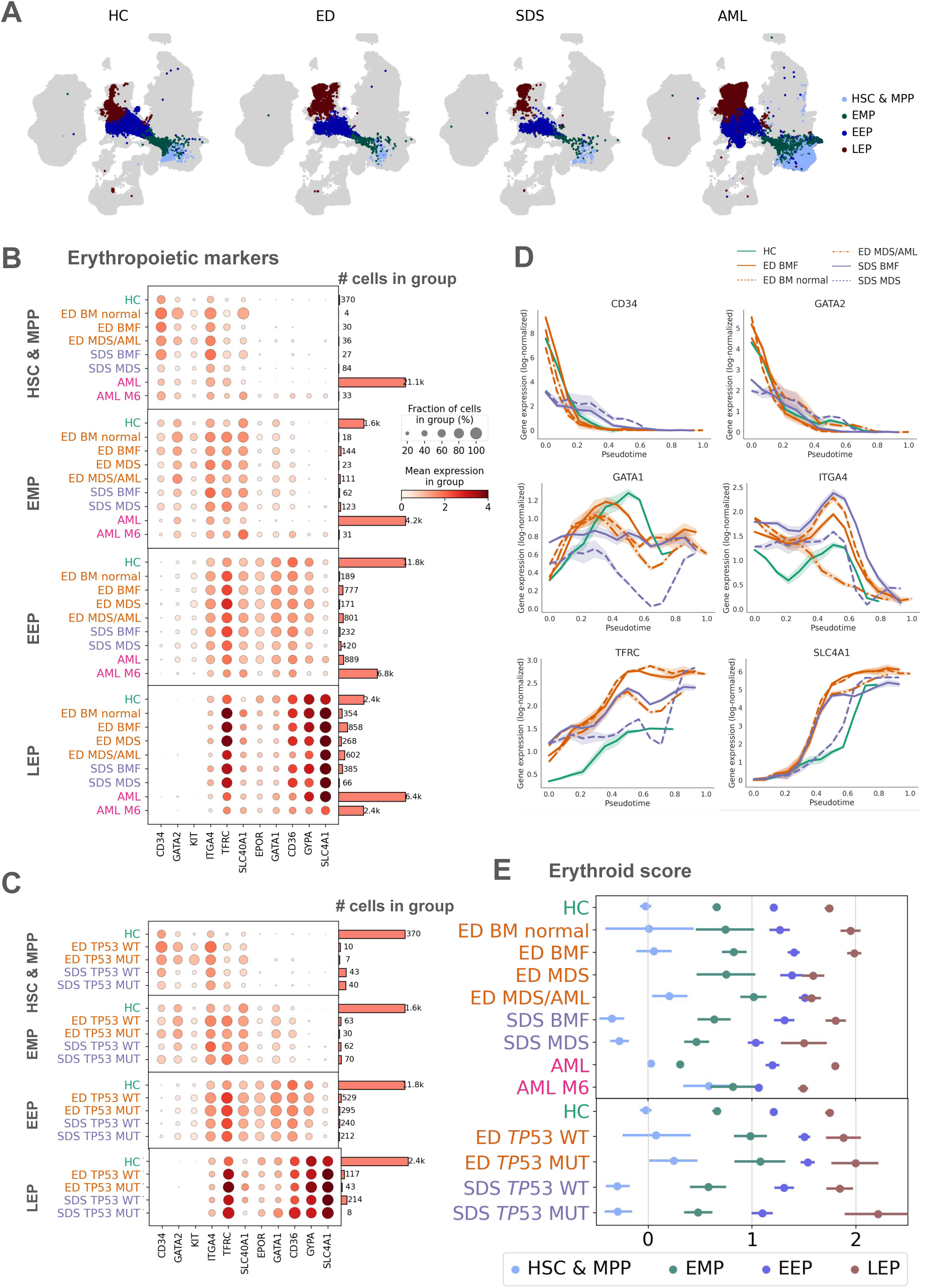
Erythropoietic cells, erythropoietic markers, and erythropoietic scores for different disease states and *TP53* mutation statuses in ED and SDS compared to healthy controls. (**A**) Erythropoietic cells highlighted (in colors) in the UMAP plot of all cells (in grey). (**B-C**) Expression of erythropoietic markers and number of cells, both across erythropoietic cell types in cells of different (**B**) disease states and (**C**) *TP53* mutation statuses. (**D**) Expression of erythropoietic markers along erythropoietic differentiation identified by pseudotime analyses. (**E**) Erythroid scores with 95% CI calculated by bootstrapping with 10,000 resamples across erythropoietic cell types in cells of different disease states and *TP53* mutation status.

Interestingly, we observed upregulation of erythroid-associated transcription measured by erythroid scores^41^ in ED but not in SDS compared to controls (Figure 3E, Supplemental Figure 11). Among EEPs, the score for ED with normal BM cellularity was lower than for all other ED disease stages (*p*<0.05, bootstrap test with 10,000 resamples, Figure 3E) and the erythroid score for the MDS/AML case was higher than in all other conditions and diagnoses even exceeding that of the AML M6 case (bootstrap *p*<0.01 and *p*<0.0001, respectively, Figure 3E, Supplemental Table 18). This pattern may reflect a compensatory upregulation of erythroid transcription under stress which is absent in the ED case with normal BM cellularity and normal blood cell counts. Divergent erythroid scores in ED and SDS point to qualitative differences in differentiation, with ED cells displaying a stronger erythroid transcriptional commitment than SDS or controls.

### ED and SDS LEPs converge on shared cell cycle and ferroptotic stress signatures that persist despite *TP53* mutation

To further investigate erythroid differentiation in ED and SDS, we analyzed cell cycle dynamics and stress-related gene expression in erythroid progenitor populations. Cell cycle scores^41^ were higher for all ED disease stages compared to control in EEPs (*p*<0.0001, bootstrap test with 10,000 resamples, Figure 4A, Supplemental Figure 11, Supplemental Table 19), indicating upregulation of cell cycle -associated genes. The distribution of cells across cell cycle phases was similar between *TP53*-mutated and wild-type cells (Figure 4B). In contrast, we detected a marked reduction in cell cycle scores in LEPs for all ED disease stages compared to control (*p*<0.0001, Figure 4A), consistent with downregulation of cell cycle transcription.

**Figure 4.**
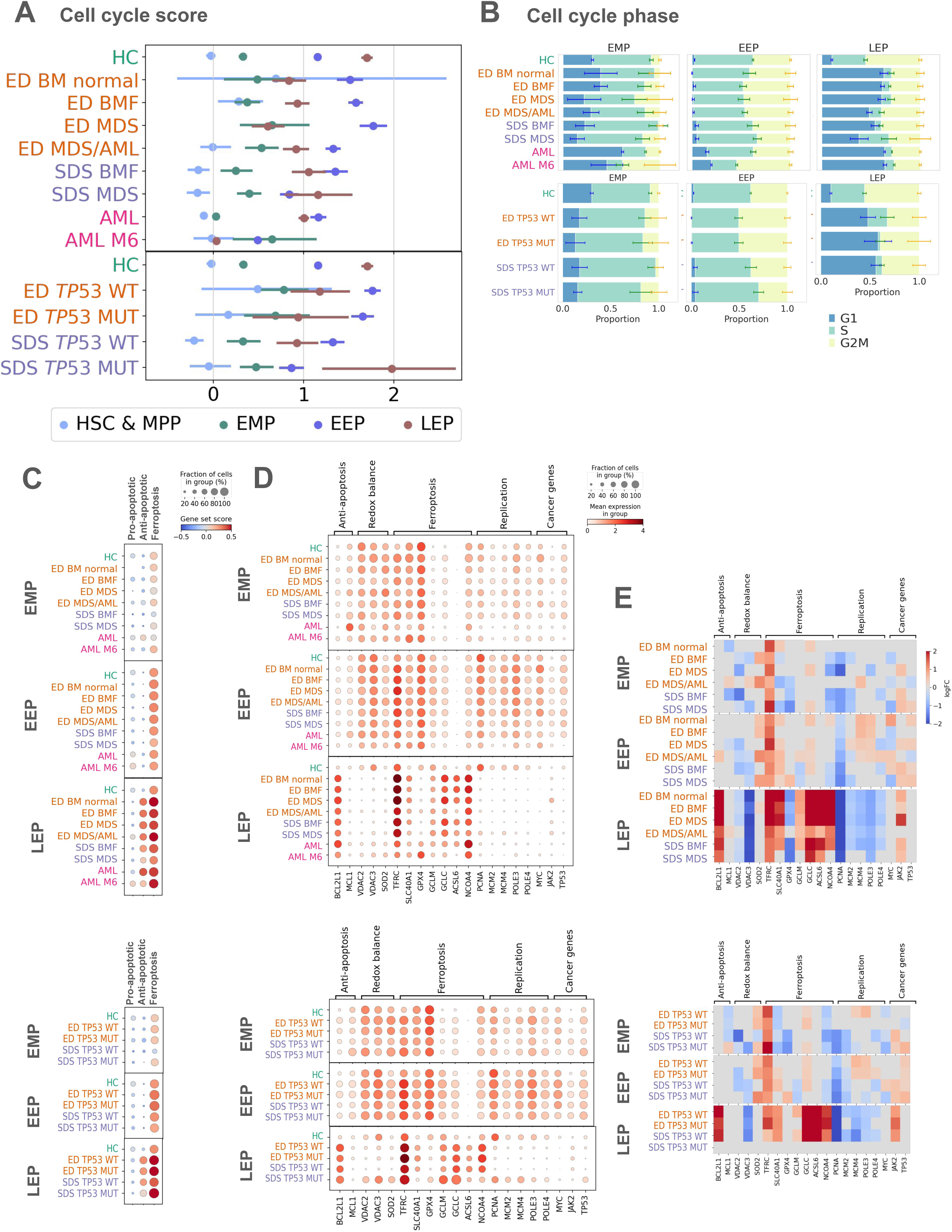
Cell cycle scores, cell cycle phase proportions, and cell death scores with their differentially expressed key genes. **(A)** Cell cycle scores (95% CI, bootstrap with 10,000 resamples) across erythropoietic cell types in cells of different disease states and *TP53* mutation status. **(B)** Cell cycle phases (95% CI, bootstrap with 10,000 resamples) across erythropoietic cell types in cells of different disease states and *TP53* mutation status. SDS *TP53*-mutated LEPs were too low in number to be included in the analysis. **(C)** Pro- and anti-apoptotic and ferroptotic gene set scores across erythropoietic cell types in cells of different disease states, and *TP53* mutation status. **(D)** Expression of key genes involved in ferroptosis, apoptosis, and replication across different disease states and *TP53* mutation status. **(E)** Heatmaps showing log_2_FC of DE results of genes presented in (D) across different disease states and *TP53* mutation status compared to healthy controls.

DE analyses further supported these findings, revealing enrichment of cell cycle–related genes in all erythropoietic cell types in both diseases (Figure 2B, Supplemental Tables 5-8). Examining cell cycle phases revealed that the proportion of S phase cells approached controls in EMPs and EEPs (45%–76% in ED and SDS, 61% in controls; and 50%–61% in ED and SDS, 59% in controls, respectively, Figure 4B, Supplemental Table 20). However, S phase cells were drastically reduced in LEPs, which resided mostly in G1 (6%–30% in ED and SDS, 34% in controls; 38%–63% in ED and SDS, 10% in controls, respectively, Figure 4B). These results suggest a change in cell cycle dynamics and erythroid differentiation in both ED and SDS, especially in LEPs with a decrease of S phase and an increase of G1 phase, a pattern reminiscent of erythroid differentiation block seen in erythroid leukemia^42^.

Given the aberrant cell cycle dynamics in LEPs and the consistently elevated expression of *TFRC*, a key marker of iron uptake and ferroptosis sensitivity, we next investigated whether dysregulated iron metabolism and replication stress might contribute to erythroid vulnerability in ED and SDS (Figure 4C). DE analyses confirmed overexpression of *TFRC* in patient cells in both diseases compared to controls throughout erythropoietic cell types (Figure 3B, Supplemental Tables 5-8). *TFRC* is required for red blood cell development and together with loss of GPX4 activity, is an important ferroptosis marker^43^. In addition to increased *TFRC* expression, we observed underexpression of *GPX4* in both ED and SDS LEPs compared to controls (Figure 4D-E). Further, *VDAC2* and *VDAC3*, which regulate mitochondrial ROS and iron flux, were downregulated in LEPs (Figure 4D-E), consistent with perturbations in iron homeostasis and redox regulation across both diseases.

Concomitant with upregulated ferroptosis, antiapoptotic genes were upregulated in both ED and SDS LEPs (Figure 4C-E, Supplemental Figure 12). Especially antiapoptotic *BCL2L1* (encoding for BCL-XL), which is downstream of *EPO* and is vital for erythropoiesis^44^, was overexpressed in ED and SDS LEPs (Figure 4D-E). In EEPs, *MCM,* and *POLE* genes that participate in replication, were overexpressed in both diseases, but their expression declined in LEPs (Figure 4D-E), potentially reflecting replication stress or failure to resolve cell cycle progression, a known driver of DNA damage and ferroptosis sensitivity^45,46^. *PCNA* was downregulated in both EEPs and LEPs in ED and SDS (Figure 4D). Its differential expression reinforces the presence of replication-associated stress, as PCNA interacts with ERCC6L2 to support replication fork progression^9^. This is consistent with ED, where ERCC6L2 is mutated, but its downregulation in SDS is nonetheless noteworthy. Moreover, we noted increased *JAK2* expression (Figure 4D-E) in LEPs, possibly reflecting adaptive changes in signaling during stressed erythropoiesis^47,48^.

Notably, the presence of *TP53* mutations in ED and SDS cells did not restore normal expression of key ferroptosis or cell cycle genes (Figure 4E, Supplemental Tables 21-24). Thus, *TP53* mutations in these contexts may be a secondary adaptation, insufficient to override the fundamental replication and redox dysregulation imposed by the primary germline *ERCC6L2* and *SBDS* deficiency. Together, these findings suggest that LEPs in ED and SDS may exist in a replication-stressed state with insufficient resolution of cell cycle progression, necessitating antiapoptotic support and predisposing cells to ferroptosis, maintained even in the presence of *TP53* mutations.

### Intrinsic differences in stem cell regulation separate ED and SDS in early hematopoiesis

To isolate fundamental changes in erythropoiesis from effects due to *TP53* mutations, cell cycle variation, sex and age, we regressed out these variables. Following this correction, HSCs, MPPs and EMPs clustered more closely for ED and controls than for SDS (Figure 5A). Differentiation trajectories showed ED and SDS converging in EEPs, and ultimately aligning closely with controls in LEPs (Figure 5A), mirroring the expression patterns described earlier (Figure 2A) and underscoring intrinsic transcriptional differences in early hematopoiesis. Stem cells are scarce in healthy BM^49^, and we detected on average six HSCs and MPPs per sample for controls (Figure 5B) and even less in ED and SDS BMF (proportion of HSCs & MPPs 0.10%, 95% CI [0.07%, 0.13%], *p*<0.0001 and 0.11%, 95% CI [0.08%, 0.15%], *p*=0.0003 respectively, compared to 0.19% in controls; Figure 5B), consistent with HSC exhaustion that leads to BMF. No HSCs and MPPs were detected in ED MDS, reflecting the characteristic hypocellular state.

**Figure 5.**
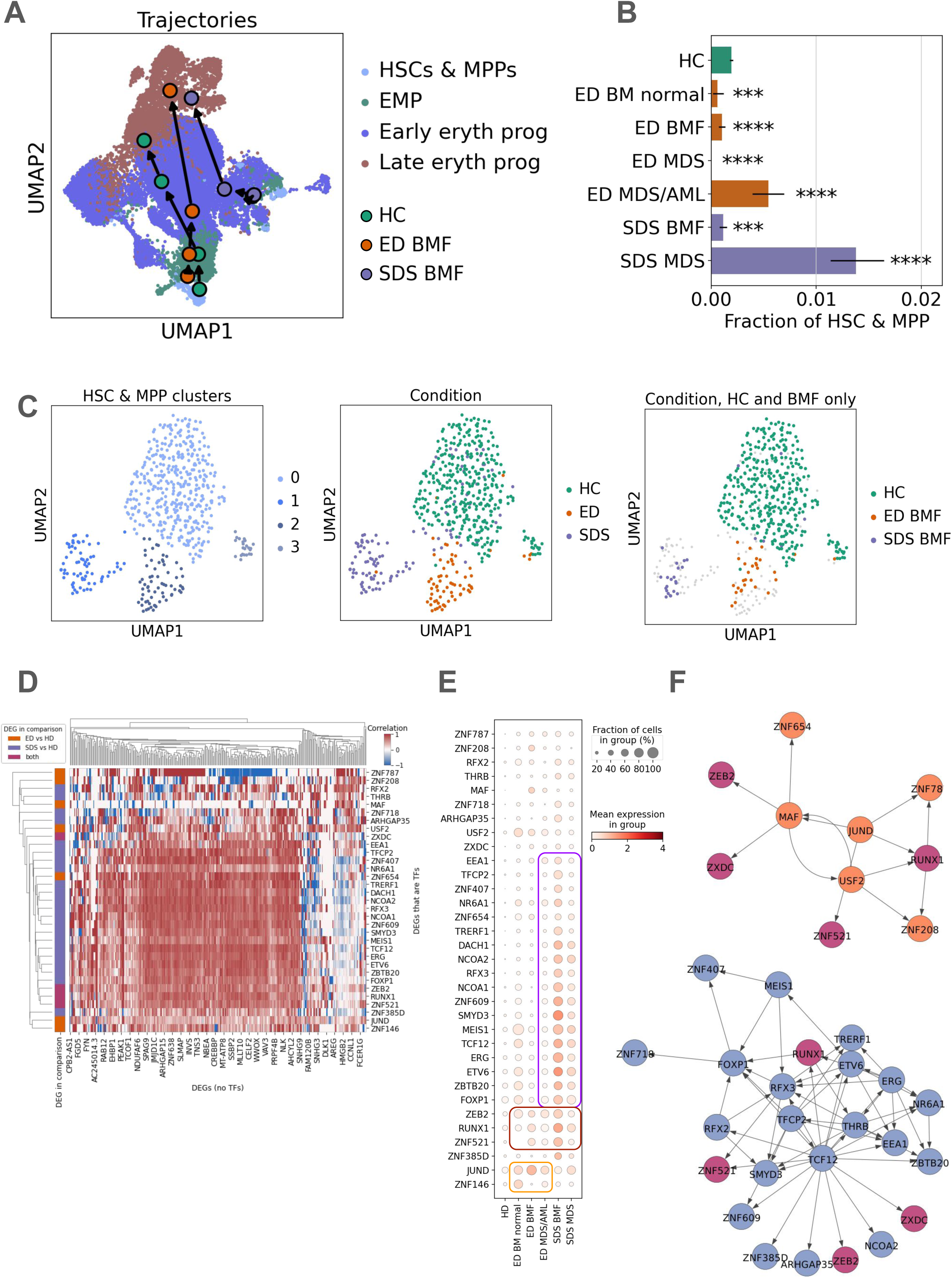
Differentiation trajectories of erythropoietic cells, proportions of HSC and MPP, and transcription factor analyses in HSC and MPP. **(A)** Differentiation trajectories of erythropoietic cells in ED BMF, SDS BMF and healthy controls depicted on a UMAP plot of these cells, after regressing out the *TP53* mutation status and cell cycle scores as well as patient/donor sex and age from gene expression levels. **(B)** Proportion of HSC and MPP in each sample group (95% CI, bootstrap with 10,000 resamples). ^∗∗∗^, *p* < 0.001; ^∗∗∗∗^, *p* < 0.0001. **(C)** HSC and MPP clusters obtained after regressing out factors listed in (A), then clustering HSC and MPP, visualized as a UMAP plot. Cells colored by clusters (left), condition (middle), and highlighting cells in ED BMF, SDS BMF, and healthy controls (right). **(D)** Heatmap showing Pearson correlation of differentially expressed TFs and non-TF DEGs. DEGs were obtained comparing HSC and MPP in ED BMF and SDS BMF to healthy controls. **(E)** Expression of differentially expressed TFs in HSC and MPP in ED BMF and SDS BMF compared to healthy controls. **(F)** Interactions of differentially expressed TFs in gene regulatory networks of HSC and MPP in ED BMF and SDS BMF.

Focused analysis of HSCs and MPPs revealed distinct clusters for ED, SDS, and controls (Figure 5C), leading us to examine whether transcription factor (TF) dysregulation underlies these early hematopoietic differences. We found partial clustering by disease, but also overlapping transcriptional changes (Figure 5D-E). SDS displayed a broader range of TF dysregulation, with a greater number of DEGs compared to ED (Figure 5D-E). To further assess the relationships between these TFs, we constructed GRNs that highlighted the differentially expressed TFs *JUND, RUNX1, USF2,* and *MAF* in ED (Figure 5F) suggesting coordinated roles in maintaining HSC self-renewal and potentially regulating early lineage priming^50–54^. Notably, *RUNX1* and *JUND* interface with *TP53* and oxidative stress pathways^55–57^, potentially contributing to erythroid-priming and antiapoptotic signaling observed in ED. In contrast, SDS cells exhibited overexpression of TFs such as *TCF12, ETV6, ERG,* and *RUNX1* (Figure 5F). These factors likely maintain HSC identity^58–60^ and could buffer against premature differentiation, rather than promote lineage skewing. Together, these findings support that ED and SDS, despite their shared phenotype of BMF, are rooted in distinct disruptions of stem and progenitor cell identity.

### *TP53*-mutated cells show erythroid differentiation block in early erythroid progenitors in ED MDS/AML patient

Finally, to understand how *TP53* mutations and chromosomal alterations contribute to leukemic transformation in ED and SDS, we examined their distribution and impact across differentiation stages. In ED BMF, *TP53* mutations accumulated only in LEPs (OR=6.5, 95% CI [1.8, 20.1], *p_adj_*=0.04, *p_adj_*>0.1 for all other cell types, Figure 6A, Supplemental Table 25). No CNVs were found in the BMF sample with the highest detected number of *TP53*-mutated cells (Figure 6B-C, Supplemental Figure 8), aligning with clinical data (Supplemental Table 2). In the ED MDS/AML case, *TP53*-mutated cells were enriched in EEPs (OR=2.8, 95% CI [2.1, 3.7], *p_adj_*=6.18×10⁻¹³), while their frequency was significantly reduced in LEPs (OR=0.4, 95% CI [0.2, 0.8], *p_adj_*=0.03; Figure 6D, Supplemental Table 25). This pattern aligns with clinical reports of erythroid-predominant AML in ED^4,5^, suggesting that transformation may involve a differentiation block specifically at the EEP-to-LEP transition, rather than a failure of early erythroid commitment. In this patient we also noticed a reduction of *TP53*-mutated cells in B and T cells (OR=0.6, 95% CI [0.3, 0.9], *p_adj_*=0.03 and OR=0.6, 95% CI [0.5, 0.8], *p_adj_*=0.003 respectively,

**Figure 6.**
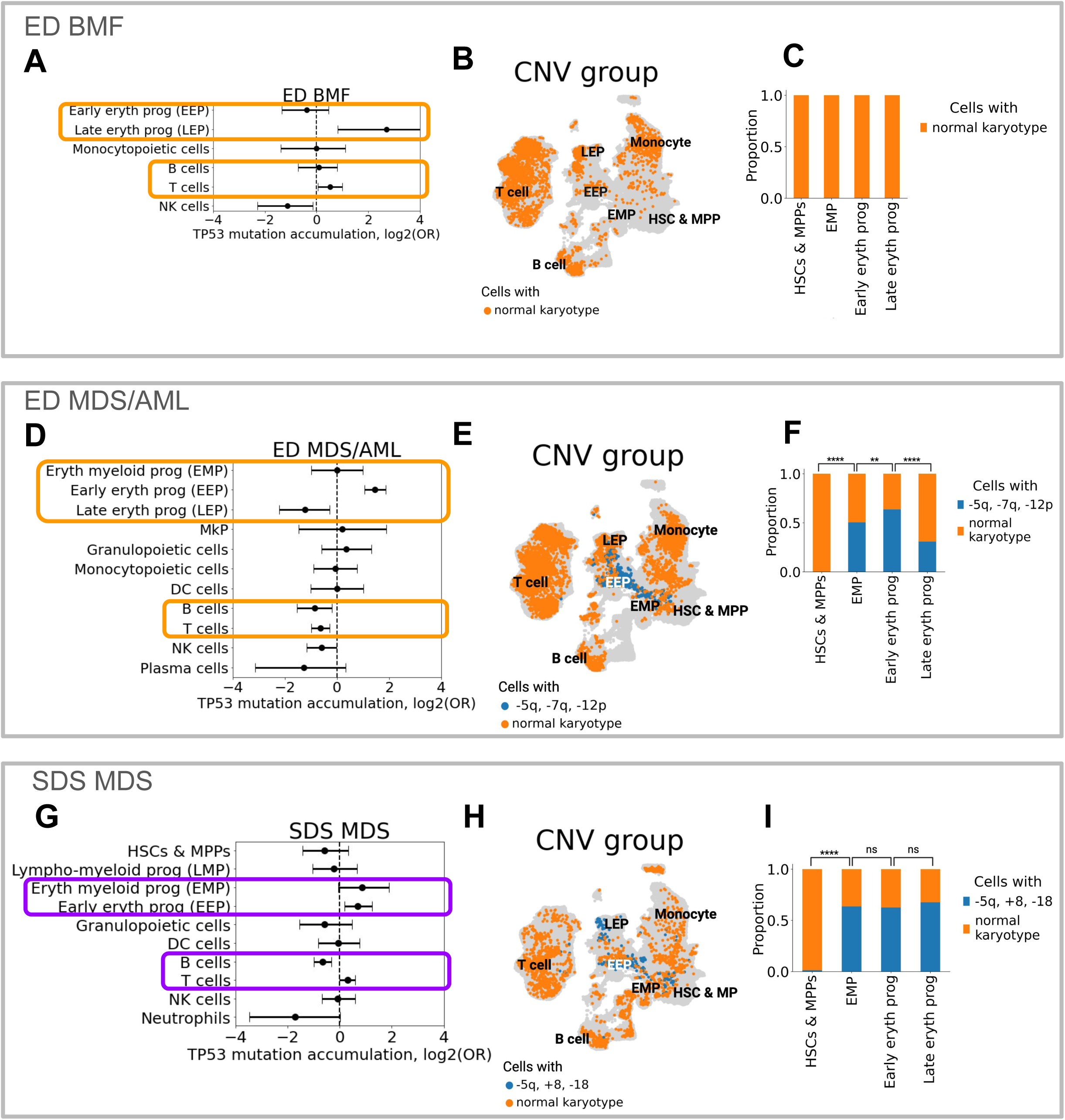
Accumulation of *TP53* mutations and chromosomal changes in two patient samples with complex karyotype: ED BMF (E2-E8a), ED MDS/AML (E10) and SDS MDS (SDS5b). (A) Accumulation and reduction of *TP53*-mutated cells in different cell types in ED BMF (n=8 samples from six patients), depicted by positive and negative log_2_(odds ratio) values, respectively. **(B)** Cells without CNVs in ED BMF (E4b) highlighted (in orange) on a UMAP of all cells (in grey). **(C)** Proportion of cells with and without CNVs per cell type in ED BMF (E4b). **(D)** Accumulation and reduction of *TP53*-mutated cells in different cell types in ED MDS/AML (n=1 sample from one patient), depicted by positive and negative log_2_(odds ratio) values, respectively. **(E)** Cells with and without CNVs in ED MDS/AML (E10) highlighted (in blue and orange, respectively) on a UMAP of all cells (in grey). **(F)** Proportion of cells with and without CNVs per cell type in ED MDS/AML (E10). **(G)** Accumulation and reduction of *TP53*-mutated cells in different cell types in SDS MDS (n=2 samples from one patient), depicted by positive and negative log_2_(OR) values, respectively. **(H)** Cells with and without CNVs in SDS MDS (SDS5b) highlighted (in blue and orange, respectively) on a UMAP of all cells (in grey). **(I)** Proportion of cells with and without CNVs per cell type in SDS MDS (SDS5b). In (A), (D) and (G) only cell types with at least five cells per cell type and TP53 mutation status are shown. Significances in (F) and (I) were calculated by a proportion test. ^∗∗^, *p*_adj_ < 0.01; ^∗∗∗∗^, *p*_adj_ < 0.0001; ns, non-significant *p*_adj_ > 0.05.

Figure 6D, Supplemental Table 25) as well as a pre-B and pro-B cell depletion (Supplemental Figure 13), which has been observed in AML^61^. In cell cycle phases, LEPs in G1 phase were reduced in the ED MDS/AML case compared to the other stages of ED (48% and 61%–63%, respectively, Figure 4B). CNV analyses locate losses in chromosomes five and seven in erythroid progenitors (Figure 6E, Supplemental Figure 14) with increased accumulation in EEPs (proportion test’s *z*=12.15, *p*=5.59x10^-34^, FDR=3.36x10^-33^ in EEPs compared to LEPs Figure 6F, Supplemental Table 26). Cells with CNVs were more often *TP53*-mutated than those without CNVs (z=26.4, *p*=4.40x10^-153,^). Together, these findings suggest that in the ED MDS/AML case, *TP53* mutations and chromosomal aberrations stall erythroid differentiation at the EEP stage, driving malignant progression by expanding a population of progenitors with genomic instability.

In the SDS MDS patient, we observed enrichment of *TP53*-mutated cells across the erythroid lineage, though only the enrichment in EEPs reached statistical significance (OR=1.6, 95% CI [1.1, 2.4], *p*_adj_=0.04, Figure 6G, Supplemental Table 25). Losses of chromosomes 5 and 18 and gains of chromosome 8 (reported in the clinical patient work-up; Supplemental Table 2) were detected throughout the erythroid lineage (Figures 6H-I, Supplemental Figure 14), contrary to the ED MDS/AML case, where the prevalence of CNVs declined in LEPs (Figure 6F). The SDS MDS patient cells with these CNVs were more often *TP53*-mutated than those without CNVs (*z*=27.0, *p*=6.75x10^-161^). LEPs in G1 phase were reduced in the SDS MDS case compared to SDS BMF (38% and 54%, respectively, Figure 4B). Together, in the SDS MDS case, TP53 mutations and CNVs are observed across the erythroid lineage, but low numbers of *TP53*-mutated cells detected in LEPs (Figure 3C) limited interpretation of their cell cycle state or potential differentiation block.

## Discussion

The hallmark of ED is a highly penetrant progression from BMF to erythroid-predominant, *TP53*-mutated myeloid malignancy. Given the limited therapeutic options and poor prognosis of *TP53*-driven leukemias^13,62,63^, identifying early disease-associated vulnerabilities in ED is crucial. To this end, we investigated the transcriptomic consequences of *ERCC6L2* deficiency focusing especially on events prior to malignant transformation with an extensive multi-tissue dataset for the rare disease.

Our findings indicate that ED cells engage maladaptive erythroid programs early in differentiation, setting them apart from SDS. Interestingly, the erythroid trajectories in ED and SDS unite over maturation: LEPs exhibited signs of ferroptotic stress, redox imbalance, and a potential dependency on *BCL2L1* in both diseases, suggesting shared vulnerabilities (Figure 7). Recognizing the emerging data on sensitivity to BCL-XL inhibition of *TP53*-mutated erythroid leukemia cells^19,64^ calls for further studies, also in the BMF context. Ferroptosis, an iron-dependent, lipid peroxidation-driven form of cell death, has been implicated in hematologic disorders including BMF syndromes^65–67^. Recent studies indicate that HSCs are particularly susceptible to ferroptosis due to their unique metabolic characteristics including iron processing^68,69^. In ribosome-related BMFs, such as SDS, disruptions in ribosomal function can lead to oxidative stress and iron accumulation, further sensitizing cells to ferroptosis^65^. In ED, ferroptosis may arise from impaired stress response capacity and redox imbalance in erythroid-biased progenitors. Hence, targeting therapeutically ferroptotic pathways via iron chelation, lipid peroxidation inhibitors, or modulation of glutathione metabolism is worth investigating to alleviate hematopoietic dysfunction.

**Figure 7.**
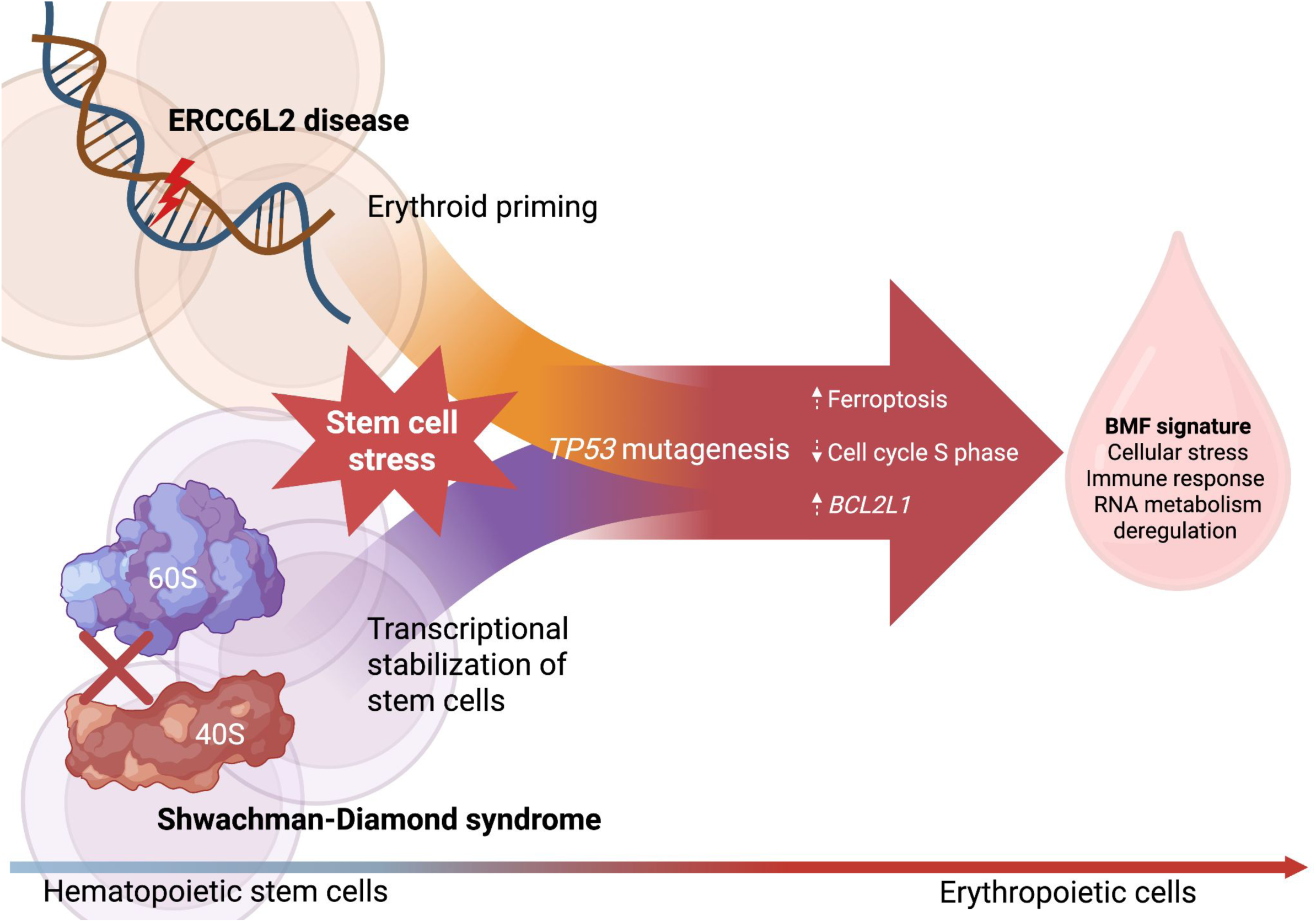
ED and SDS show distinct stem cell identity converging into shared erythroid stress. Overall, our results show that while ED and SDS diverge in early hematopoietic disruption, they converge into a shared BMF phenotype driven by erythroid stress that consists of ferroptosis, cell cycle S phase reduction, and prominent *BCL2L1* overexpression. Image created with BioRender.

Consistent with stress pathway dysregulation, we identified gene regulatory networks indicated in DNA damage and oxidative stress (involving, *e.g.*, *RUNX1, JUND*), undermining the cellular capacity to buffer against DNA damage and contributing to HSC exhaustion. In SDS, core HSC regulators seem to preserve progenitor identity under ribosomal stress. High-confidence TF interactions such as *TCF12-RUNX1* and *ETV6-MEIS1* point to compensatory mechanisms that stabilize progenitor programs and support multipotency in a hypoproliferative, translationally stressed environment. As *RUNX1* is deeply embedded in TF circuits with *ERG, ETV6, MEIS1, FOXP1* in SDS, changes in *RUNX1* expression could propagate through these networks, amplifying effects on hematopoietic stability, especially under chronic dysfunction. The differential expression of *RUNX1* emerged as a shared regulatory node in ED and SDS, potentially integrating stress signals and modulating downstream programs in both diseases. Despite distinct root causes of these diseases, both germline defects ultimately impose a bottleneck on erythropoiesis, channeling hematopoiesis into a shared, stressed erythroid transcriptome.

In ED, *TP53*-mutated clones arise in stem and progenitor cells, supporting the notion that *TP53* mutations provide a selective advantage under chronic stress by sustaining BM cellularity, similar to the adaptations seen in SDS^11,12^. However, this adaptation appears inherently unstable, as biallelic *TP53* inactivation represents a pivotal step toward leukemic transformation. Interestingly, we did not see differences in stress signatures between *TP53*-mutated and wild-type cells neither in ED nor SDS. This underscores the biological complexity of hematopoietic failure. While p53 inactivation is often thought to alleviate differentiation blocks by disabling DNA damage checkpoints^70,71^, our findings do not support a substantial rescue effect at least in the erythroid lineage, where *TP53*-mutated ED cells retained prominent stress signatures.

As a limitation of our study, we could not distinguish between monoallelic and biallelic *TP53* mutations due to recognized challenges in genotyping genes with low expression^23,24^. Biobanking of well-characterized samples, paired with advanced single-cell, genomic, and proteomic tools, will be critical for future studies to uncover the root causes of leukemogenesis in ED.

Our findings reinforce the importance of studying primary patient material. Patient-derived samples reveal disease-relevant mechanisms, such as retained *ERCC6L2* transcripts, or the stepwise accumulation of *TP53* mutations, that may not emerge in knockout models. They also allow interrogation of heterogeneous cell states across disease stages.

In conclusion, while ED and SDS diverge in early hematopoiesis, they converge into a shared BMF phenotype driven by erythroid stress that consists of ferroptosis, cell cycle S phase reduction, and prominent *BCL2L1* overexpression. This study provides the first patient-level single-cell map of ED and SDS as a resource for future work on these diseases and *TP53*-driven leukemogenesis.

## Data Availability

Data may be obtained from the authors upon reasonable request when compatible with European General Data Protection Regulation (GDPR) and with permission from the University of Helsinki.

## Acknowledgements

We thank Anna Näätänen and Emma Saarinen for sample processing and technical help, Anna Kuosmanen, Kristen Nader, and Sofia Forstén for discussions on scRNA-seq data analyses, Bulat Zagidullin for technical assistance on GPU usage, Merja Heinäniemi for locating the HCA data, Samuele Canellieri for discussions on GRN analyses, and Minna Eriksson for technical assistance. Single-cell RNA sequencing and single-cell amplicon sequencing was performed at the Institute for Molecular Medicine Finland (FIMM) Single-Cell Analytics and Genomics units supported by HiLIFE and Biocenter Finland (BF). RNA sequencing was performed at Biomedicum Functional Genomics Unit (FuGU). The authors wish to acknowledge the IT Center for Science (CSC), Finland, and the Institute for Molecular Medicine Technology Center (FIMM TC) for generous computational resources.

This work was supported by Research Council of Finland (#349760, #322675), iCAN Digital Precision Cancer Medicine Flagship, Sigrid Jusélius Foundation, Cancer Foundation Finland, and Special governmental grant for health sciences and research. This project was also supported by an unrestricted educational grant by Incyte Biosciences and grants from Instrumentarium Science Foundation, Orion Research Foundation, Biomedicum Helsinki Foundation, Päivikki and Sakari Sohlberg Foundation, Emil Aaltonen Foundation (S.P.M.D.), K. Albin Johansson Foundation (I.K.), Paulo Foundation, Ida Montin Foundation, Finnish Hematology Association, Blood Disease Research Foundation (I.K. and S.P.M.D.), and iCANDOC doctoral education pilot in precision cancer medicine (H.N.). O.K. is a K. Albin Johansson Cancer Research Fellow, Foundation for the Finnish Cancer Institute. T.S. received expert fees from Amgen, Abbvie, GSK, Otsuka Pharma, Celgene, Jansen-Cilag unrelated to this work. C.A.H. received research funding from BMS/Celgene, Kronos Bio, Novartis, Oncopeptides, Orion Pharma, WntResearch, and the Innovative Medicines Initiative 2 project HARMONY, and expert fees from Amgen and Autolus unrelated to this work. U.W.-K. received expert fees from Amgen and Incyte unrelated to this work.

I.K. and S.P.M.D are PhD candidates at the University of Helsinki and would like to thank the Doctoral Programs for Integrative Life Science and Biomedicine, respectively, for salary support. This work is submitted in partial fulfillment of the requirement for the PhD.

## Authorship Contributions

L.L., I.K., S.P.M.D., J.L., U.W.-K., E.P., and O.K. designed the study; M.H., R.N., S.K., T.S., and U.W.-K. collected the samples and clinical data; I.K., I.I., L.K and T.R. performed cell culture and sample processing; L.L., J.L. and E.P. designed and performed single cell genotyping; S.A., M.V-K., and C.A.H. provided AML sample data; L.L., I.K., S.P.M.D., H.N., I.I., J.K. and K.M. performed bioinformatic analyses and created the figures; L.L., I.K., S.P.M.D., H.N., I.I., J.K., U.W.-K., E.P., and O.K. interpreted the data; L.L. and I.K. wrote the manuscript; L.L., I.K., S.P.M.D., L.K., U.W.-K., E.P. and O.K. revised the manuscript; all authors approved the final version of the manuscript.

