## Supplemental Figures for "Distinct Stem Cell Identities Converge into Shared Erythroid Stress in ERCC6L2 Disease and Shwachman-Diamond Syndrome"

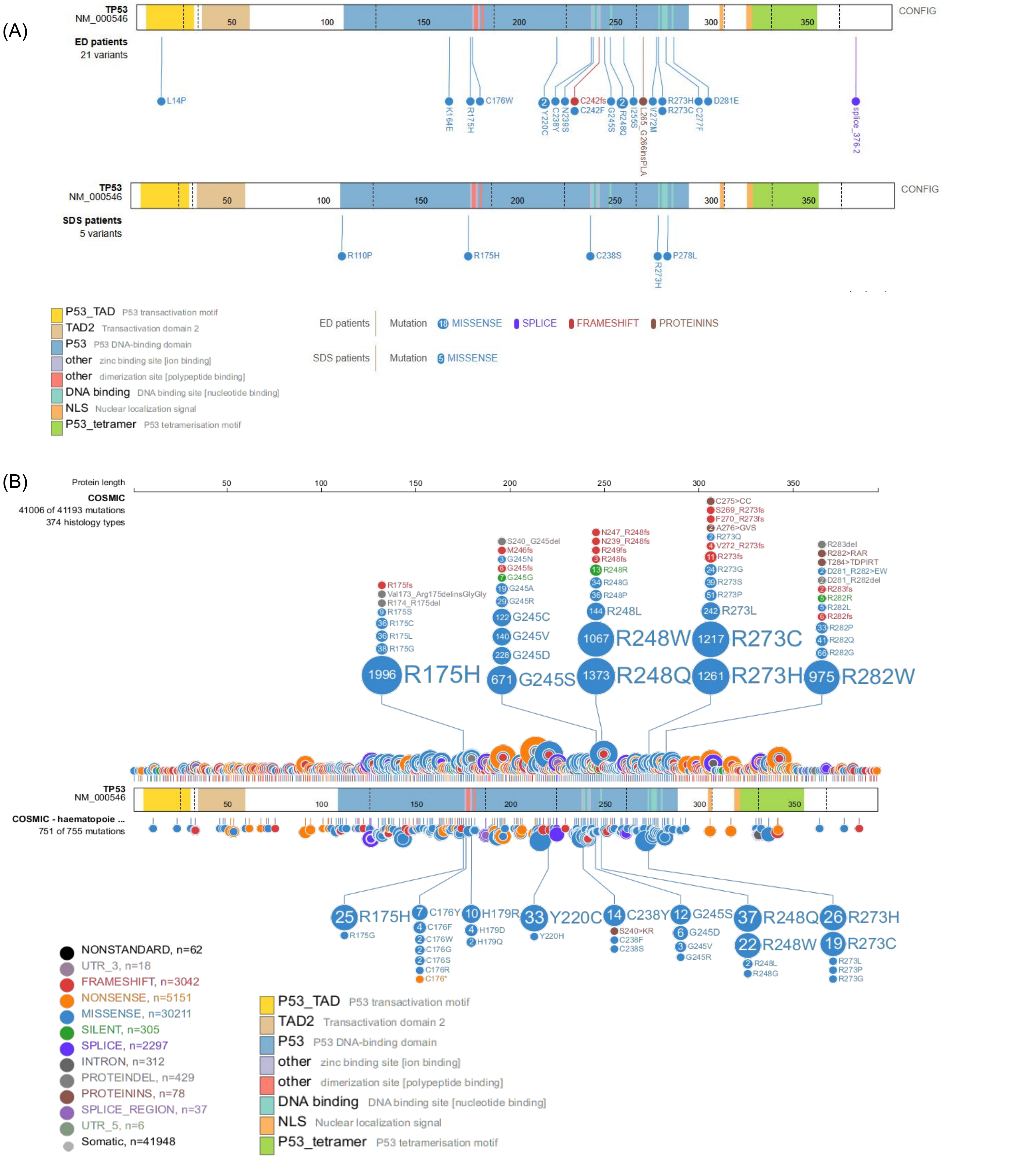


**Supplemental Figure 1. LOLLIPOPS. (A-B)** Lollipop plots of somatic *TP53* variants in ED and SDS patients included in **(A)** the scRNA-seq data, and **(B)** in the Catalogue Of Somatic Mutations In Cancer (COSMIC) and its subset of haematopoietic neoplasms fibroblasts. ED patient *TP53* variants N239S (c.761A>G) and L265_G266insPLA (c.795_796insCCTCTTGCT) are not yet in ClinVar and therefore their positions were specified by hand.

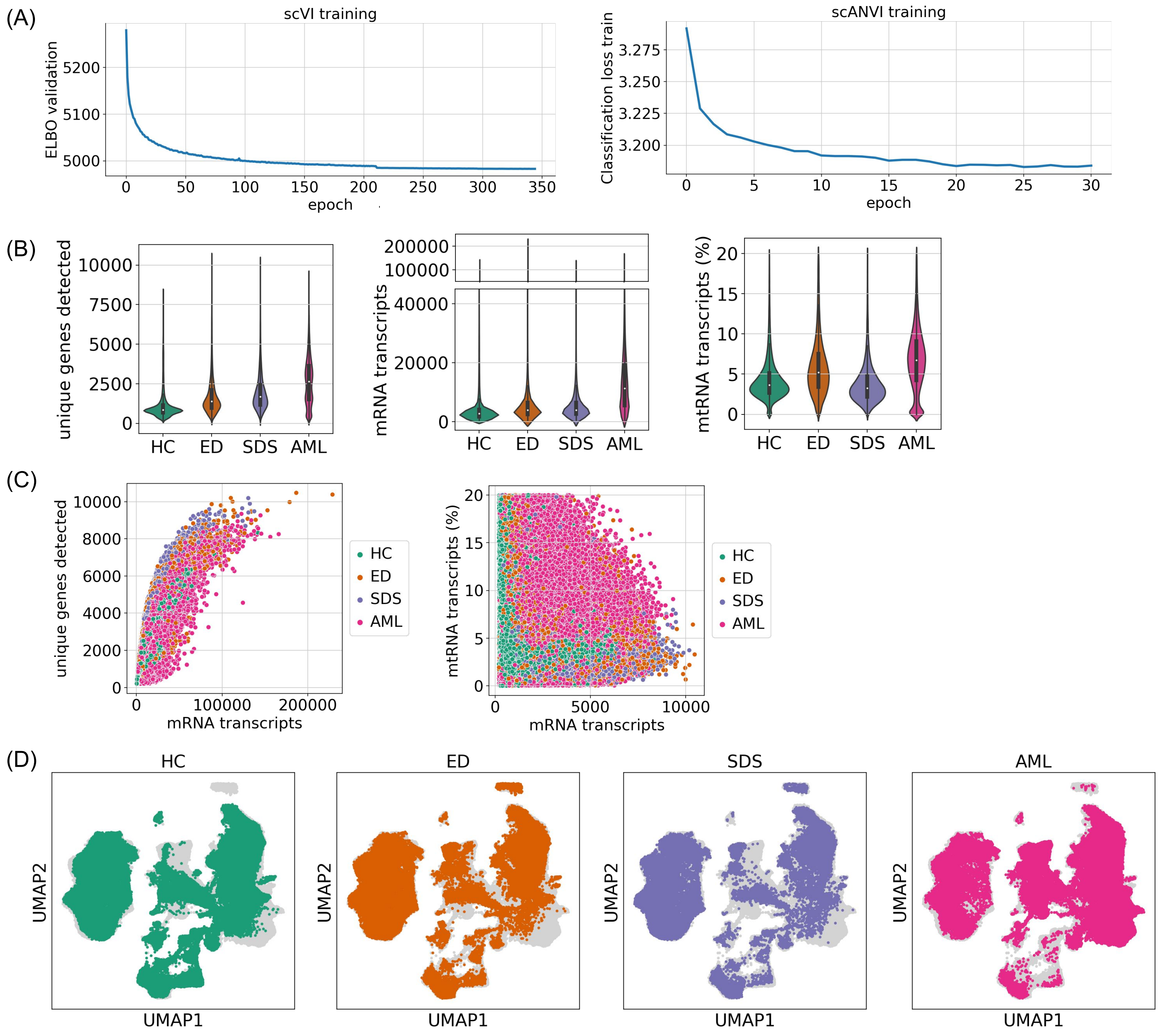


**Supplemental Figure 2. scRNA-seq data integration and QC. (A)** ELBO validation of scVI model training and classification loss of scANVI model training. **(B)** Number of unique genes detected per condition, number of mRNA transcripts per condition, and percentage of mtRNA transcripts per condition. **(C)** Number of mRNA transcripts plotted against number of unique genes detected, and number of unique genes detected plotted against percentage of mtRNA transcripts. **(D)** UMAP plots highlighting the cells of each condition (in color) on top of all cells (in grey).

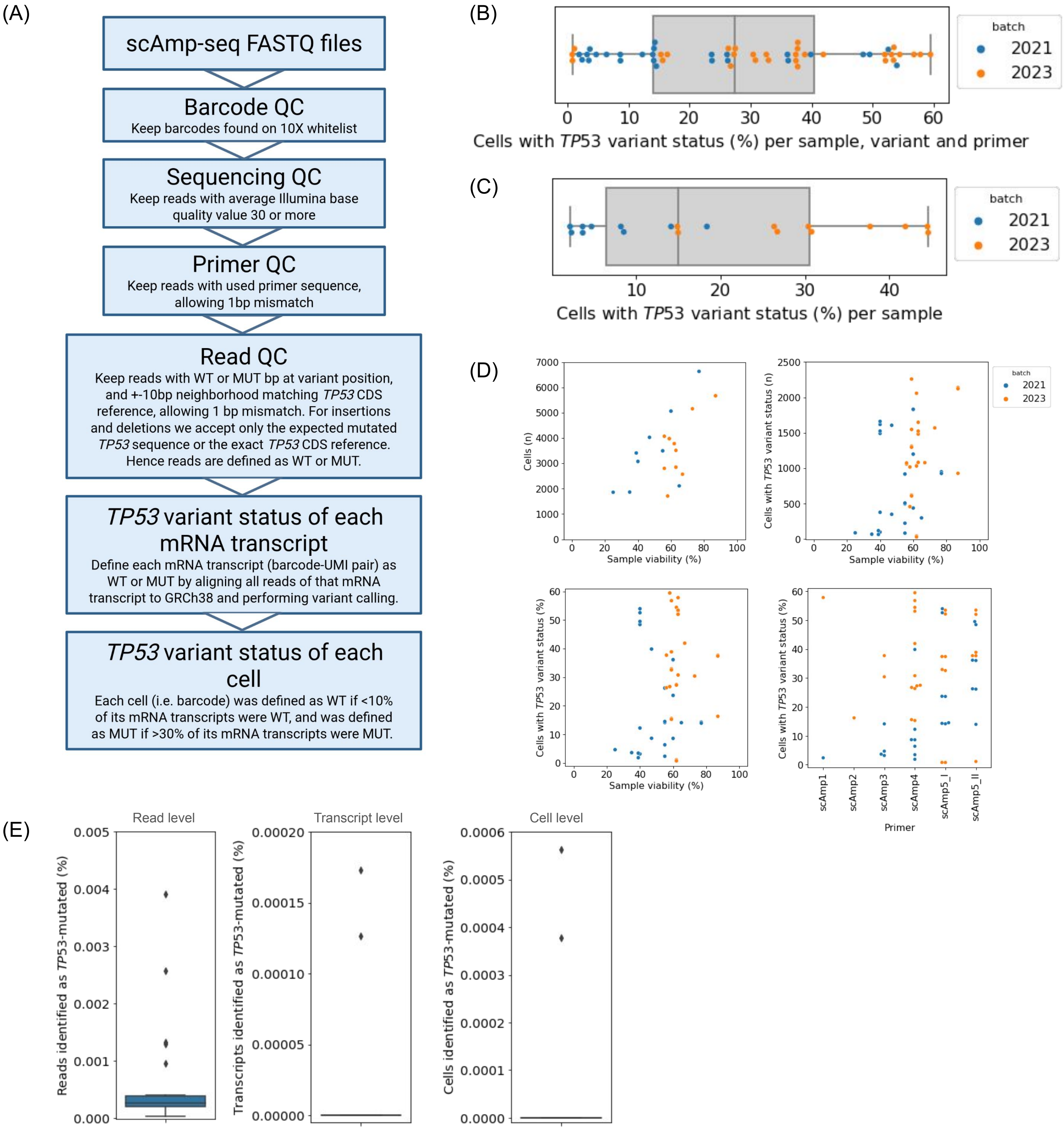


**Supplemental Figure 3. *TP53* variant status identification pipeline and QC. (A)** Overview of *TP53* variant status identification pipeline. **(B-C)** The percentage of cells for which the *TP53* variant status was identified **(B)** for each sample and each primer and each *TP53* variant for which the sample was analyzed, and **(C)** for each sample over all *TP53* variants for which the sample was analyzed. **(D)** Percentage of viable cells in sample counted before proceeding with 10X Genomics protocol, compared to the number of cells per sample in scRNA-seq after QC (upper left panel), compared to the number of cells and percentage of cells per sample and per *TP53* variant for which the *TP53* variant status was identified (upper right and lower left panel, respectively), and compared to the percentage of cells per sample and per *TP53* variant for which the *TP53* variant status was identified for each primer separately (lower right panel). **(E)** Testing our pipeline on *TP53* wild-type positions showed that the pipeline identified few false positives at the read level, and almost no false positives at the mRNA transcript and cell level.

(A)

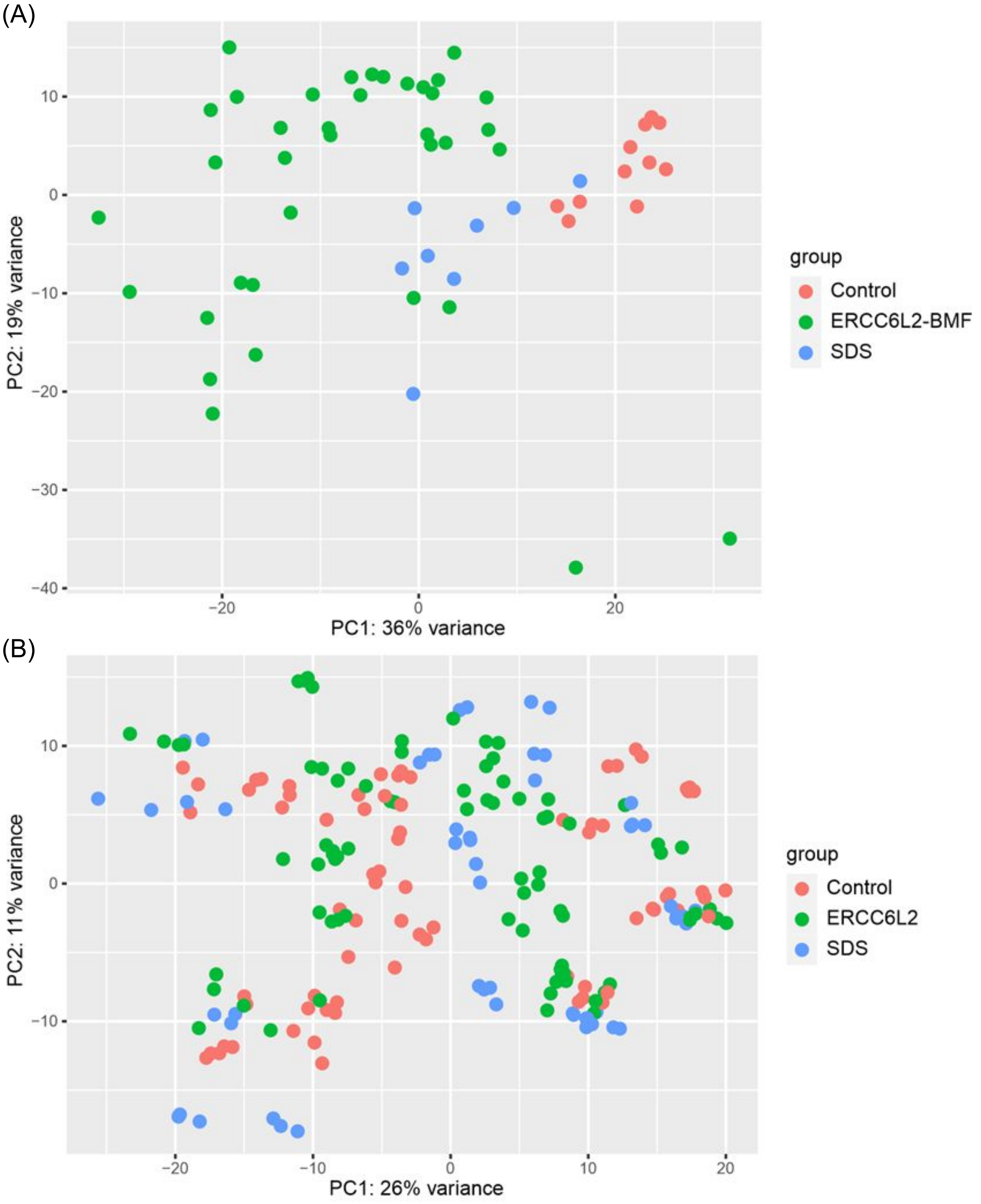


(B)



**Supplemental Figure 4. Blood and fibroblast RNA-seq QC. (A-B)** PCA plots showing sample variance in **(A)** blood and **(B)** fibroblasts.

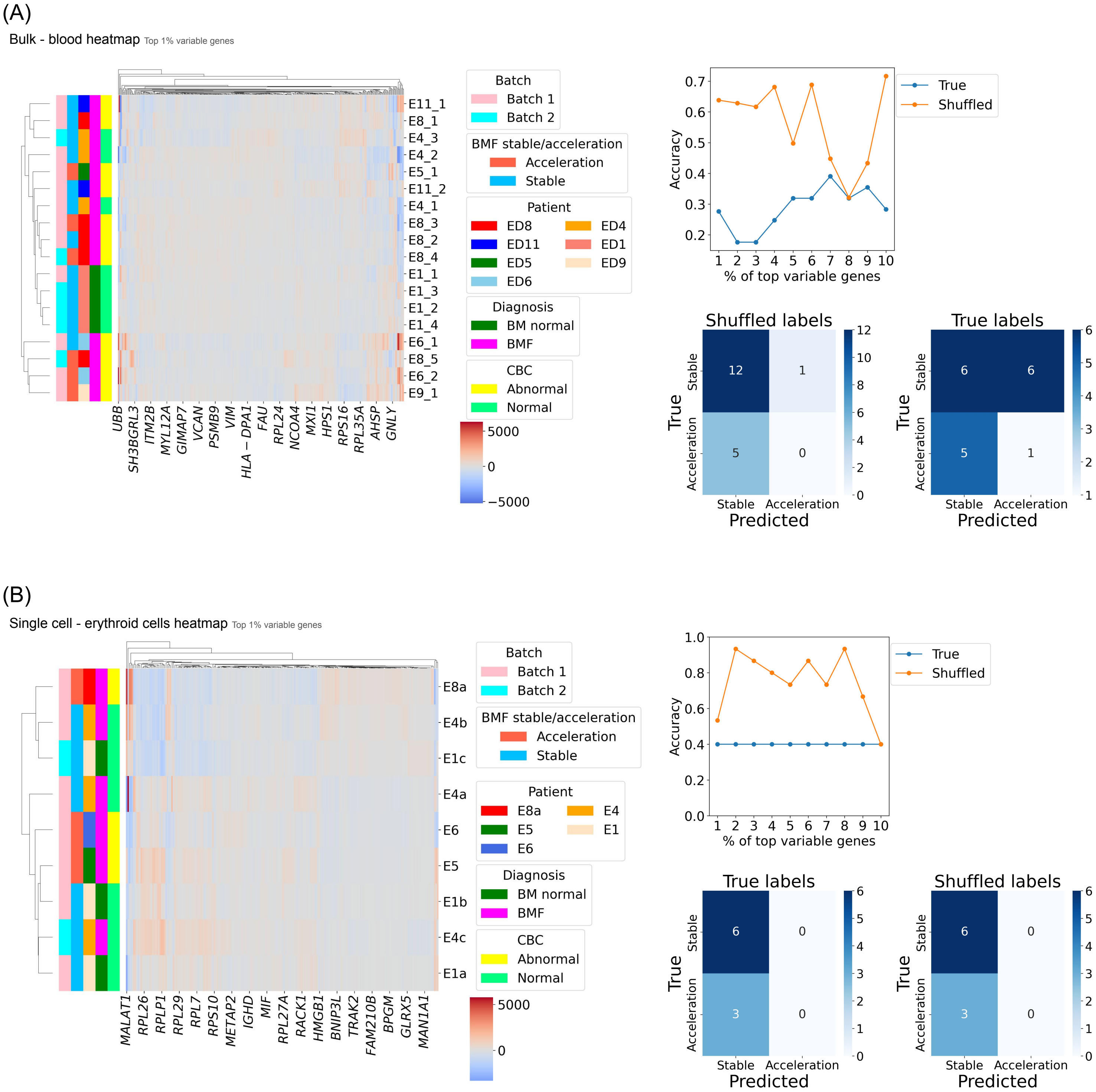


**Supplemental Figure 5. Gradient boosting on BMF stable/acceleration status. (A)** Clustering of the most variable genes (1%) based on their normalized expression between the samples in blood (left panel), the accuracy of the XGBoost model on true and shuffled labels (upper right), and the confusion matrices showing the XGBoost results predicted with top 1% most variable genes. (lower right). **(B)** Clustering of the most variable genes (1%) based on their normalized expression between the samples in BM using erythroid cells only (left), the accuracy of the XGBoost model on true and shuffle labels (upper right), and the confusion matrices showing the XGBoost results predicted with top 1% most variable genes (lower right). In the accuracy plots, the X-axis shows the percentage of the most variable genes used for prediction.

(A)

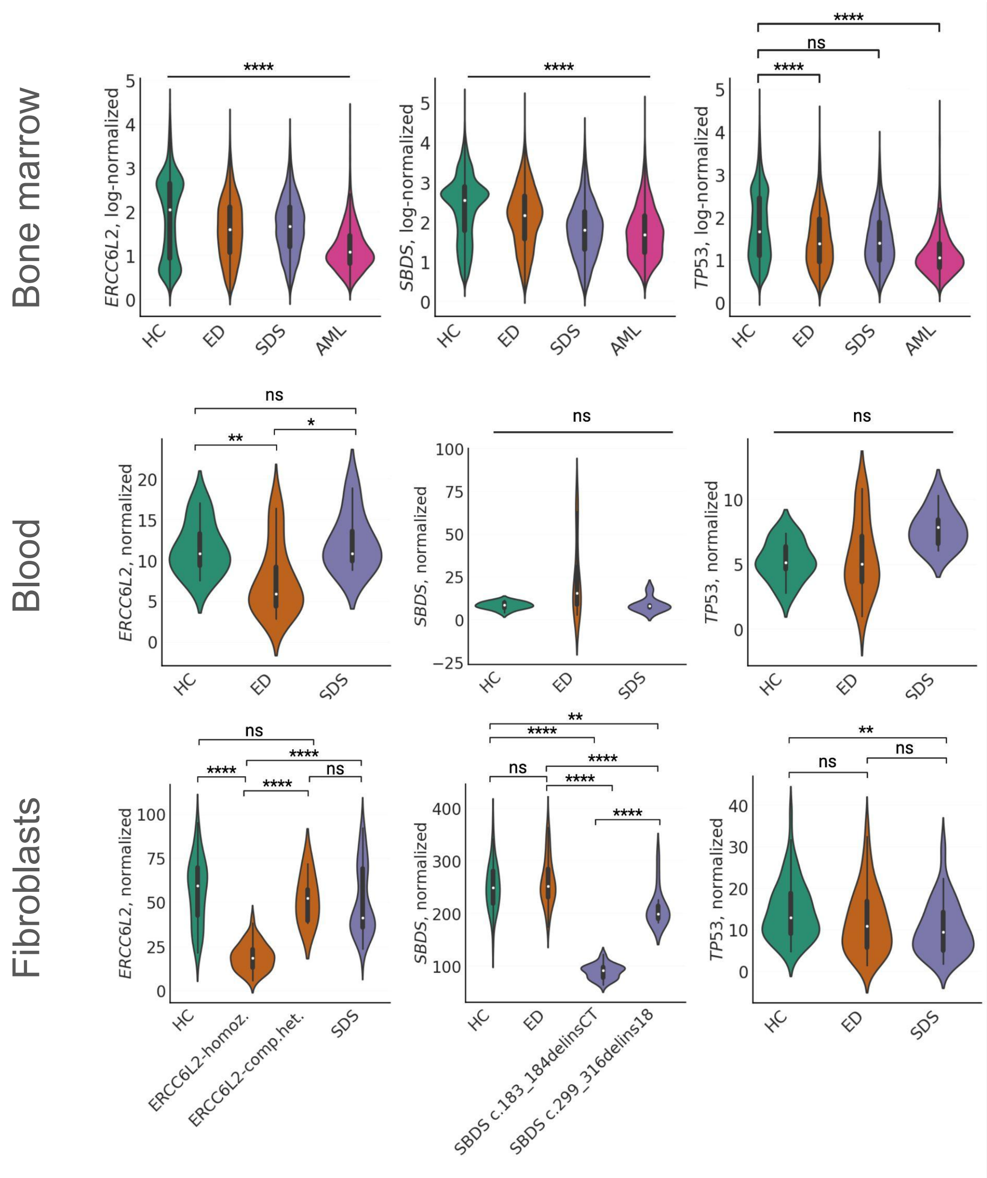


(B)

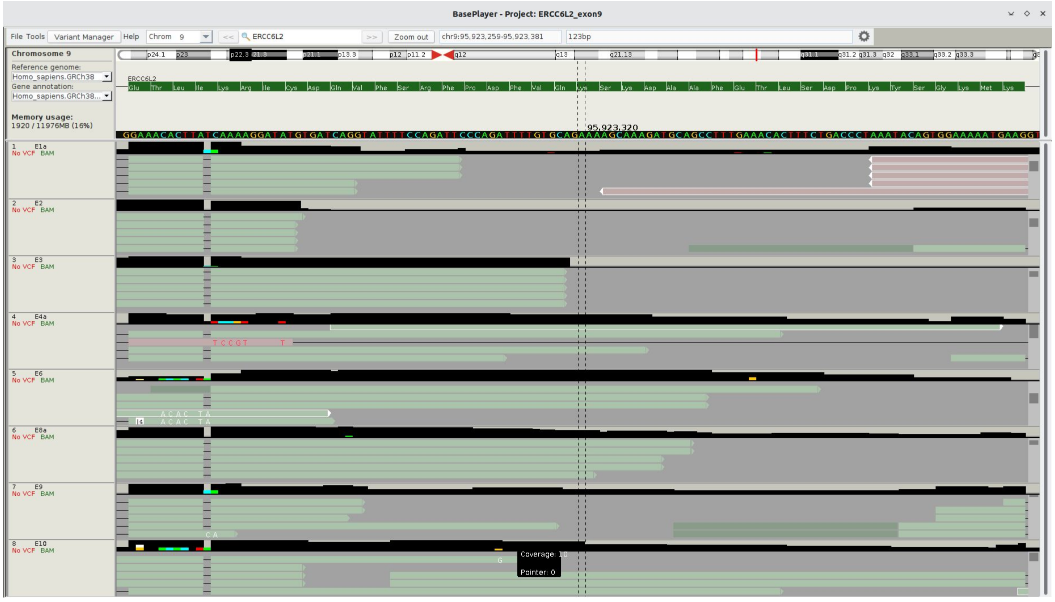


**Supplemental Figure 6. *ERCC6L2*, *SBDS*, and *TP53* expression in bone marrow, blood, and fibroblasts.(A)** Expression of *ERCC6L2*, *SBDS*, and *TP53* in bone marrow (top), blood (middle), and fibroblasts (bottom). One-way ANOVA with Tukey's test: *, $p_{adj} < 0.05$; **, $p_{adj} < 0.01$; ***, $p_{adj} < 0.001$; ****, $p_{adj} < 0.0001$; ns, not significant. The flat bar indicates that all pairwise comparisons between the groups are statistically significant at the given level (bone marrow) or non significant (blood). **(B)** scRNA-seq reads aligned to *ERCC6L2* reference using BasePlayer show the single nucleotide deletion in exon 9 caused by the *ERCC6L2* germline variant c.1424delT (p.Ile475ThrfsTer36, rs768081343).

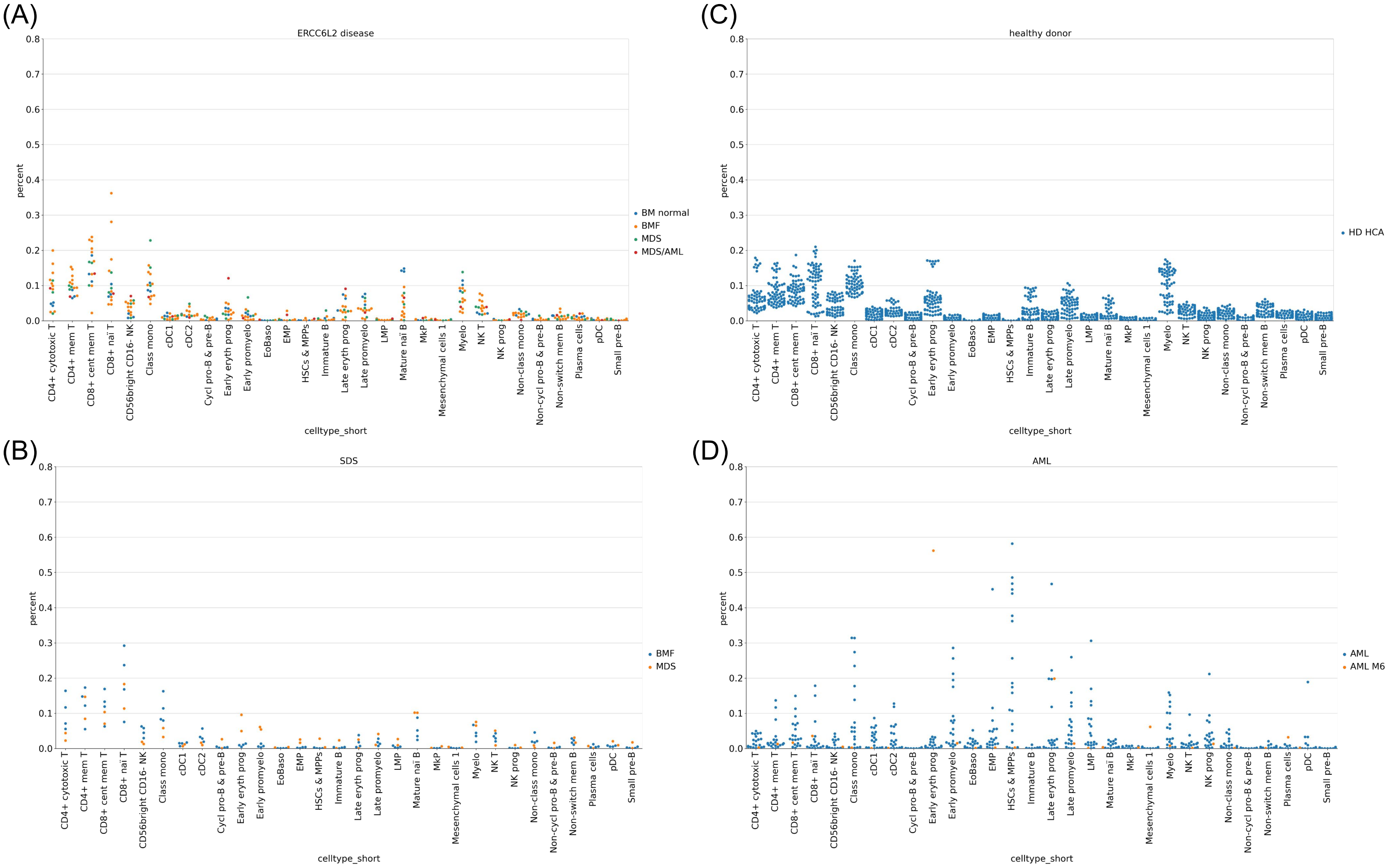


**Supplemental Figure 7. Cell type distributions in BM.** Cell type distribution plots showing the percentage of cells per cell type in **(A)** ED, **(B)** SDS, **(C)** HC, and **(D)** AML for each disease stage.

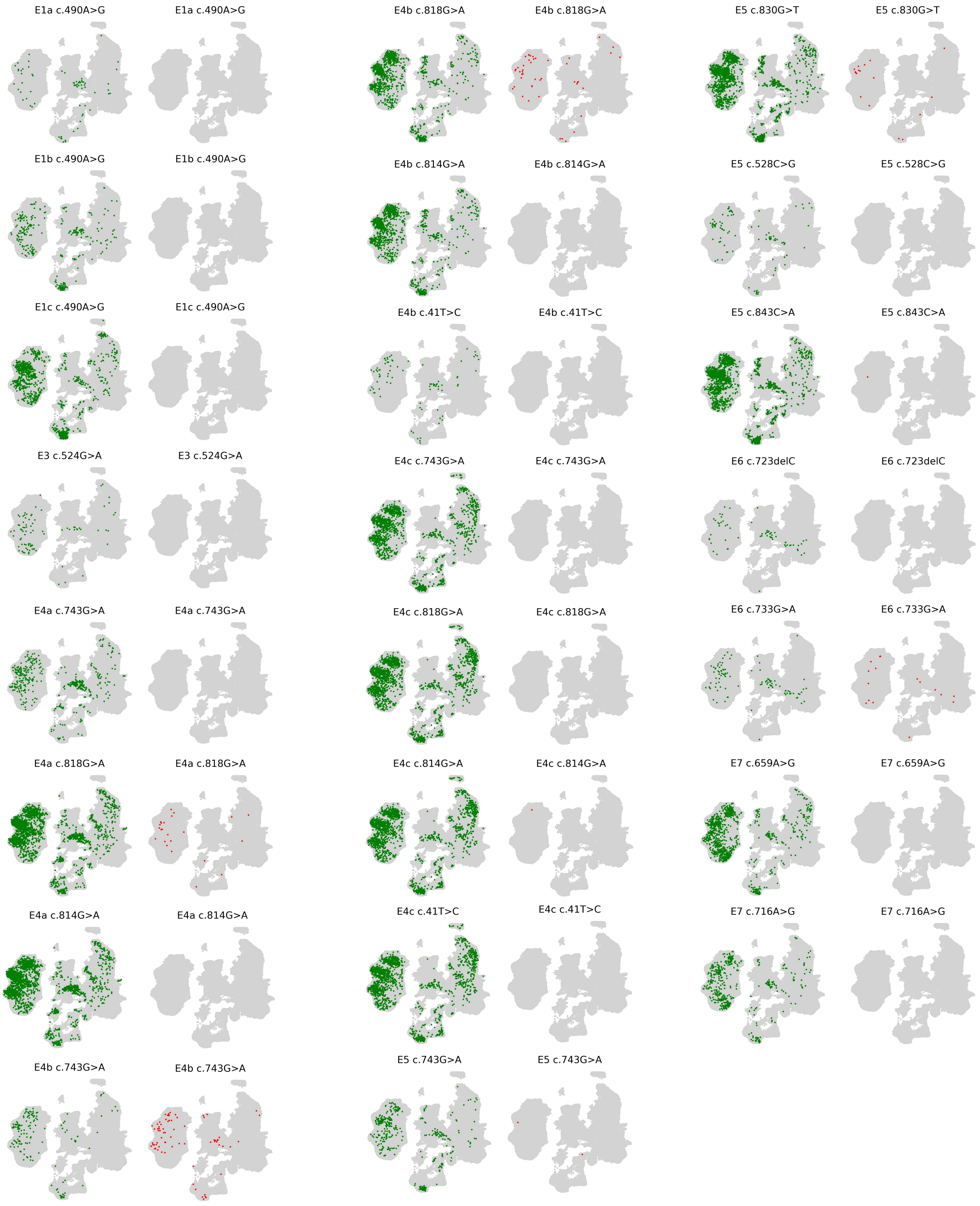


**Supplemental Figure 8. *TP53* wild-type and mutated cells per sample and *TP53* variant in ED patients E1-E7.** UMAP highlighting *TP53* wild-type (green) and mutated (red) cells per sample and *TP53* variant.

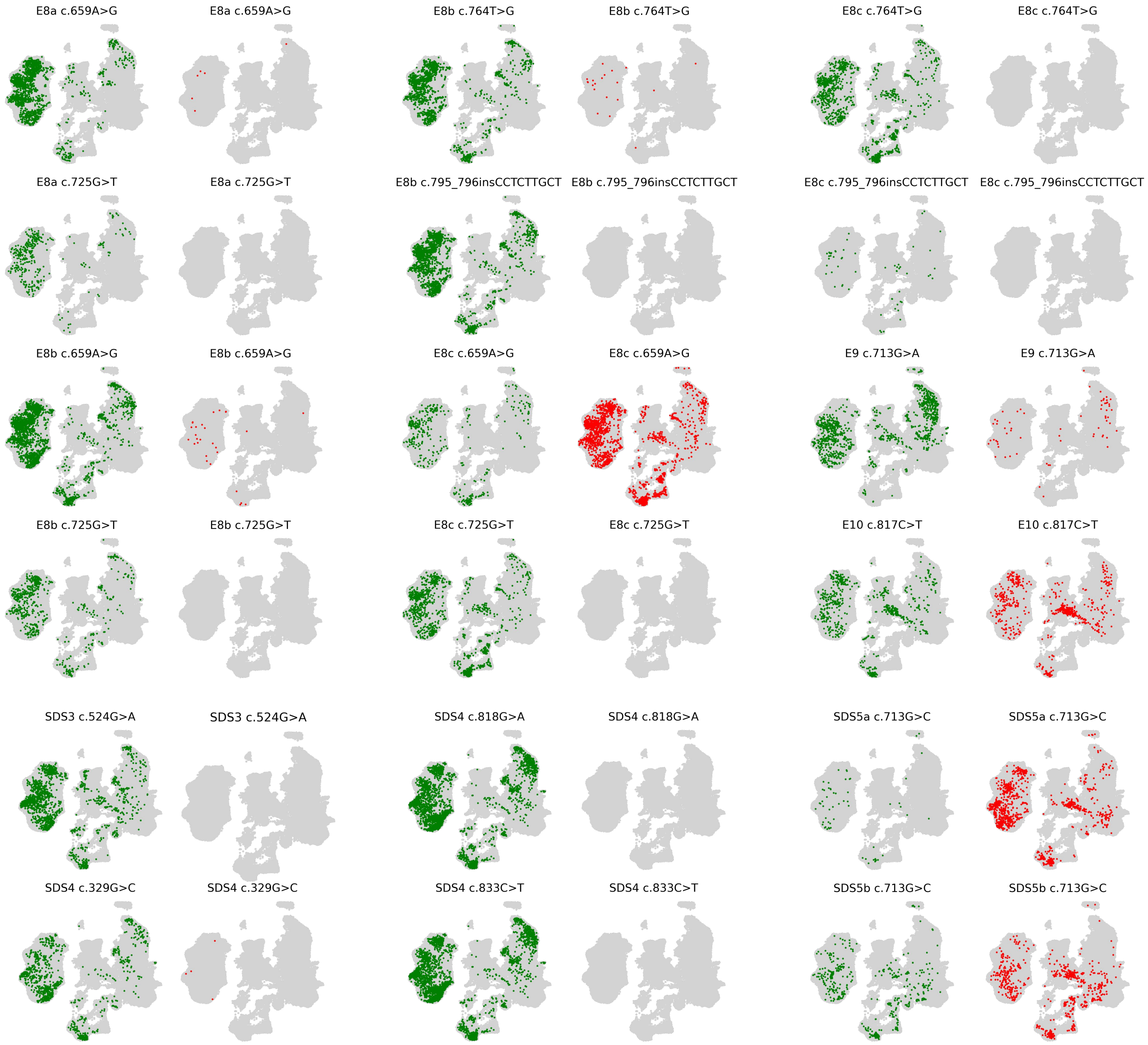


**Supplemental Figure 9. *TP53* wild-type and mutated cells per sample and *TP53* variant in ED patients E8-E10 and SDS.** UMAP highlighting *TP53* wild-type (green) and mutated (red) cells per sample and *TP53* variant.

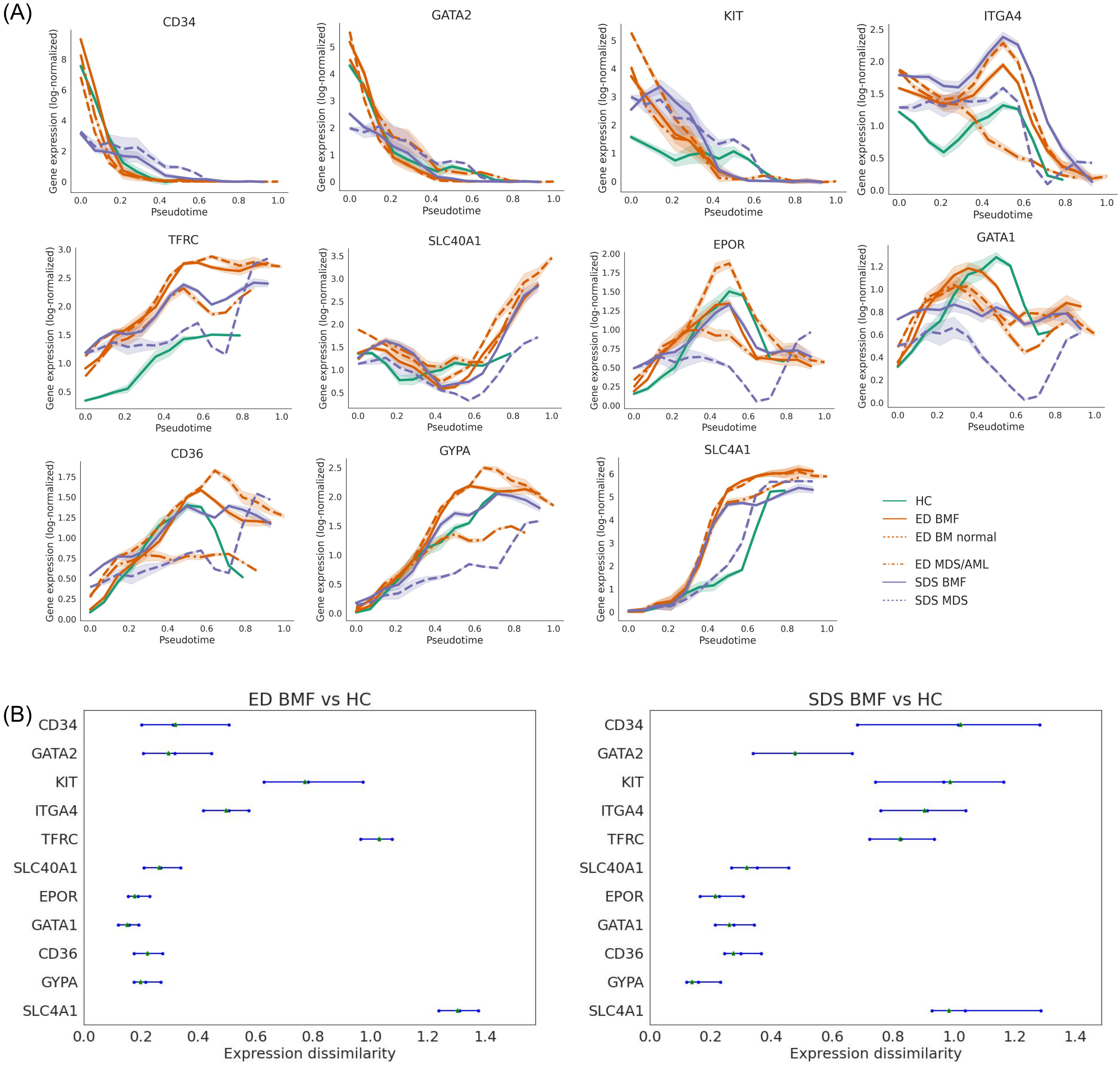


**Supplemental Figure 10. Expression of key erythroid markers along pseudotime and dissimilarity between them. (A)** Expression of key erythroid markers along pseudotime in ED, SDS and HC. **(B)** Expression dissimilarity was measured by mean absolute distance between expression along pseudotime for ED BMF vs HC (left panel) and SDS BMF vs HC (right).

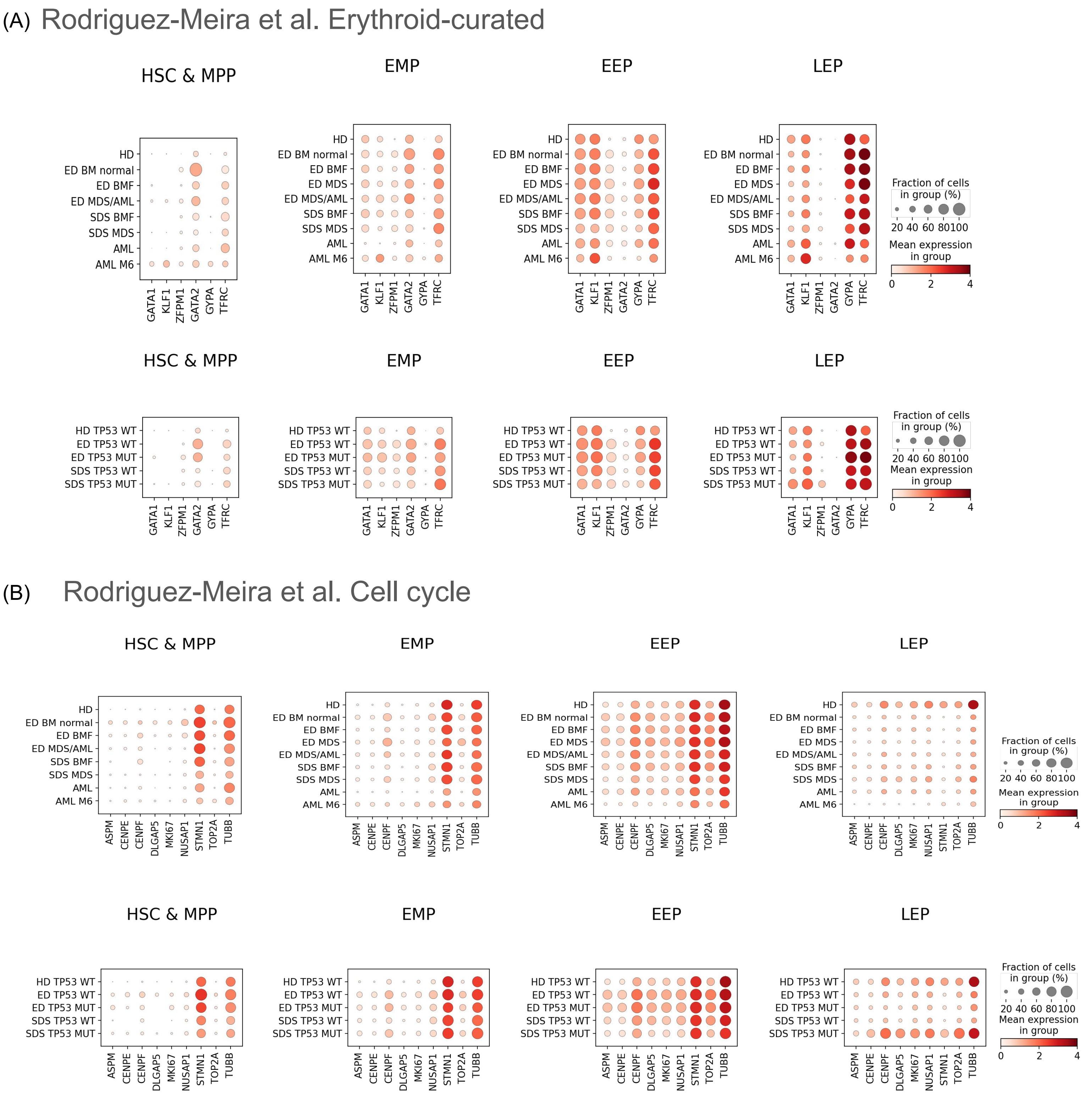


**Supplemental Figure 11. Dot plot visualization of curated gene expression signatures across disease states (top) and *TP53* mutation status (bottom) in erythroid cells. (A)** Erythroid differentiation genes from Rodriguez-Meira et al.[34] erythroid curated list and **(B)** cell cycle -related genes from Rodriguez-Meira et al.[34].

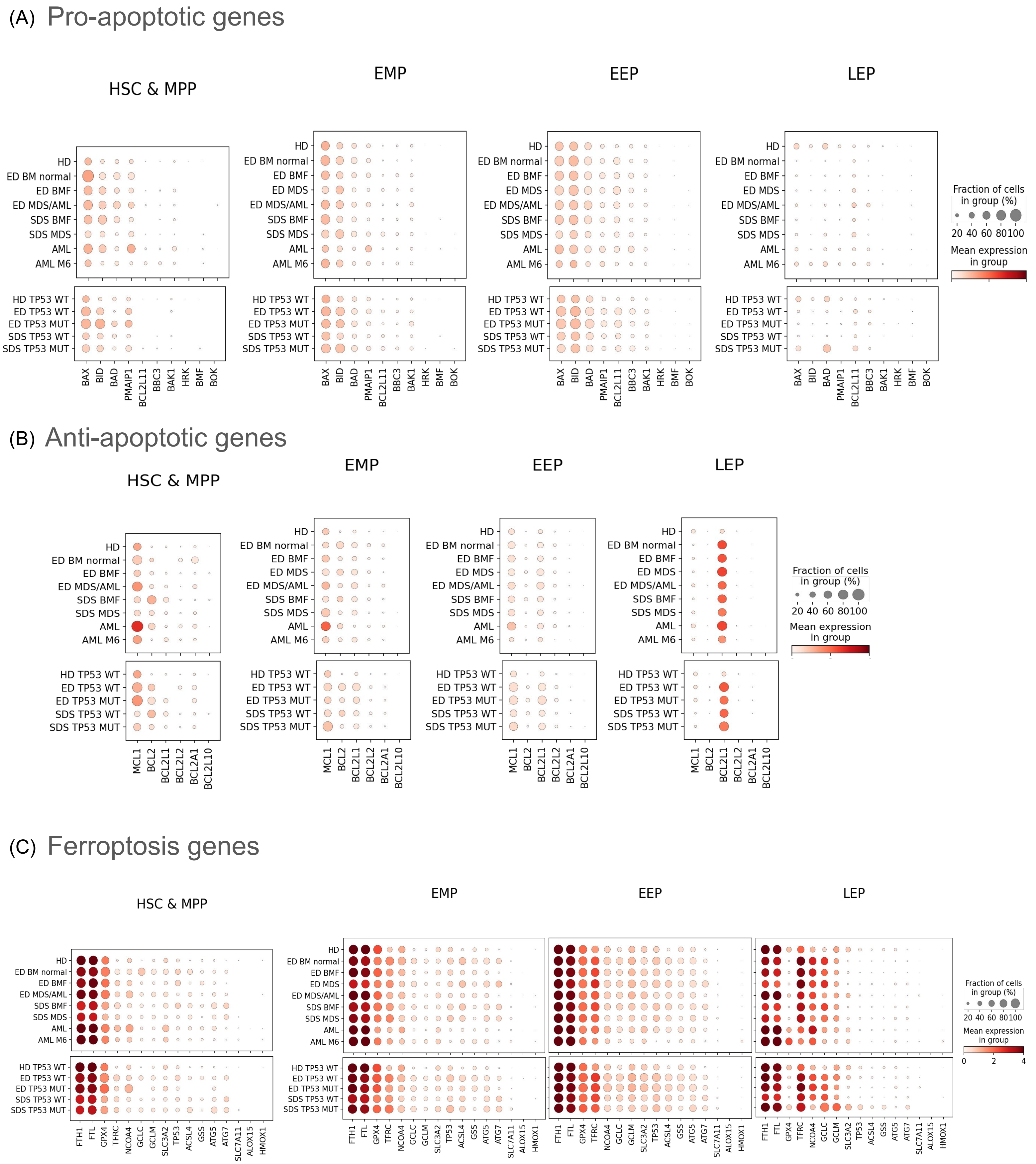


**Supplemental Figure 12. Dot plot visualization of curated gene expression signatures across disease states and TP53 mutation status in erythroid cells. (A)** Pro-apoptotic genes adapted from Ferrarini et al.[49] **(B)** Anti-apoptotic genes adapted from Ferrarini et al.[49] **(C)** Ferroptosis-related genes derived from KEGG[51] and Jiang et al.[50].

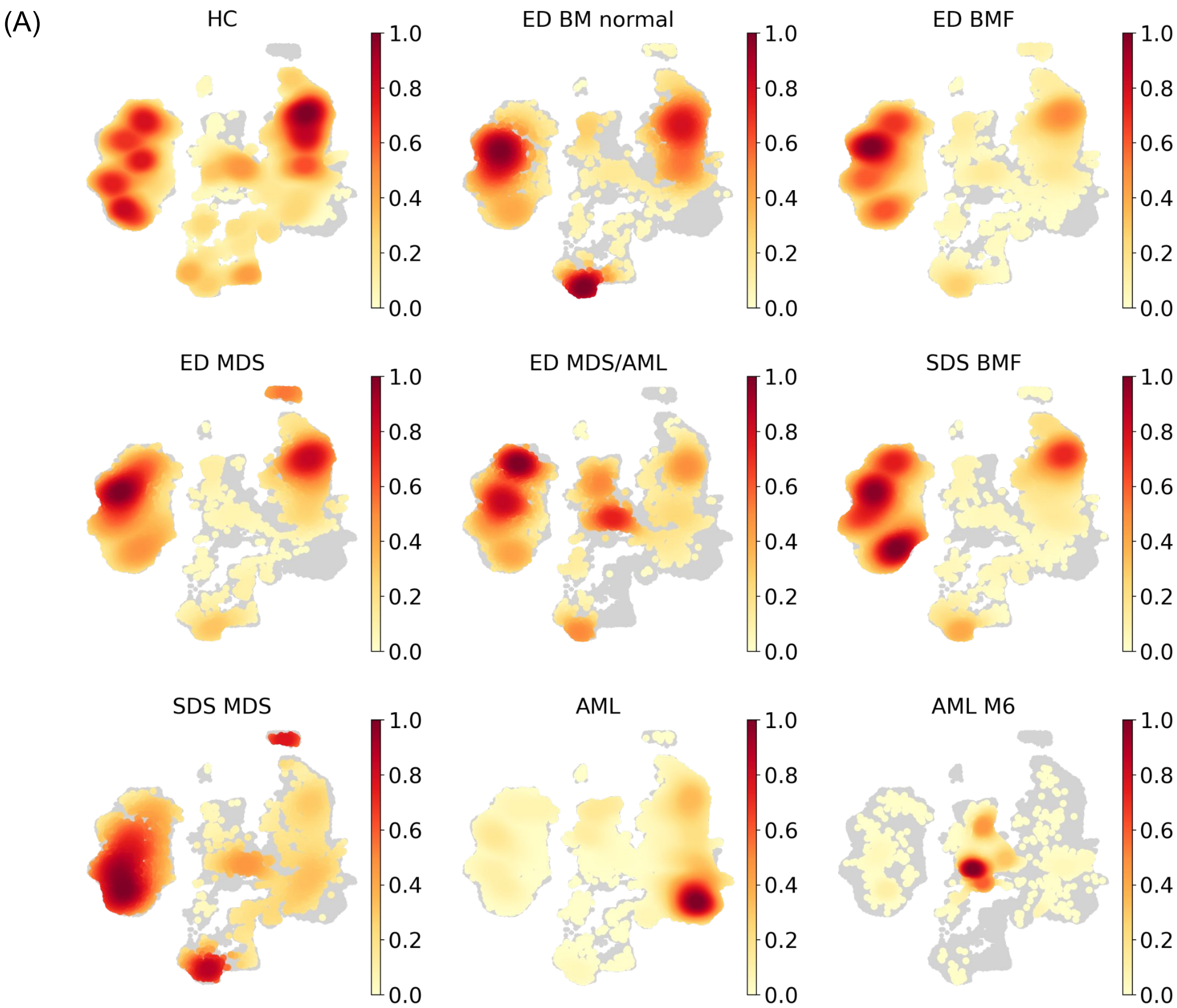


**Supplemental Figure 13. Density plots. (A)** Gaussian kernel density estimates scaled to be between zero and one per condition/diagnosis depicting the relative amount of cells in HC, ED, SDS and AML.

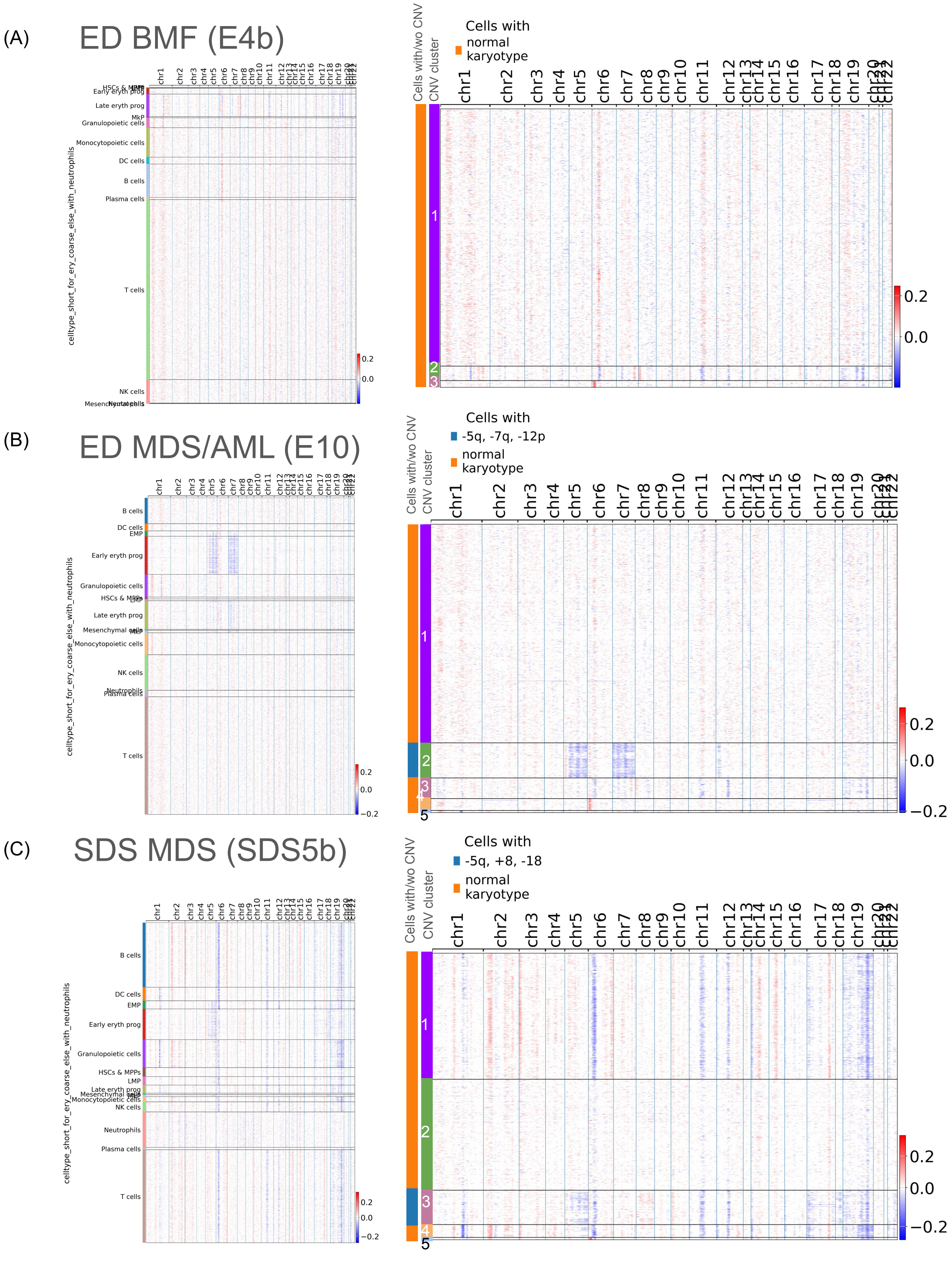


**Supplemental Figure 14. CNV profiles across cell types and clusters in ED MDS/AML case and SDS MDS case. (A-B)** CNV heatmap for **(A)** ED BMF (E4b) patient, **(B)** ED MDS/AML (E10) patient, and **(C)** SDS MDS (SDS5b) patient showing chromosomal gains and losses across the genome. Cells are grouped by annotated cell types (left panel) and CNV profile (right panel). Each row represents a single cell; each column corresponds to a genomic position ordered by chromosome. Color scale indicates relative CNV signal reflecting inferred copy number deviations from baseline (gains in red, losses in blue).

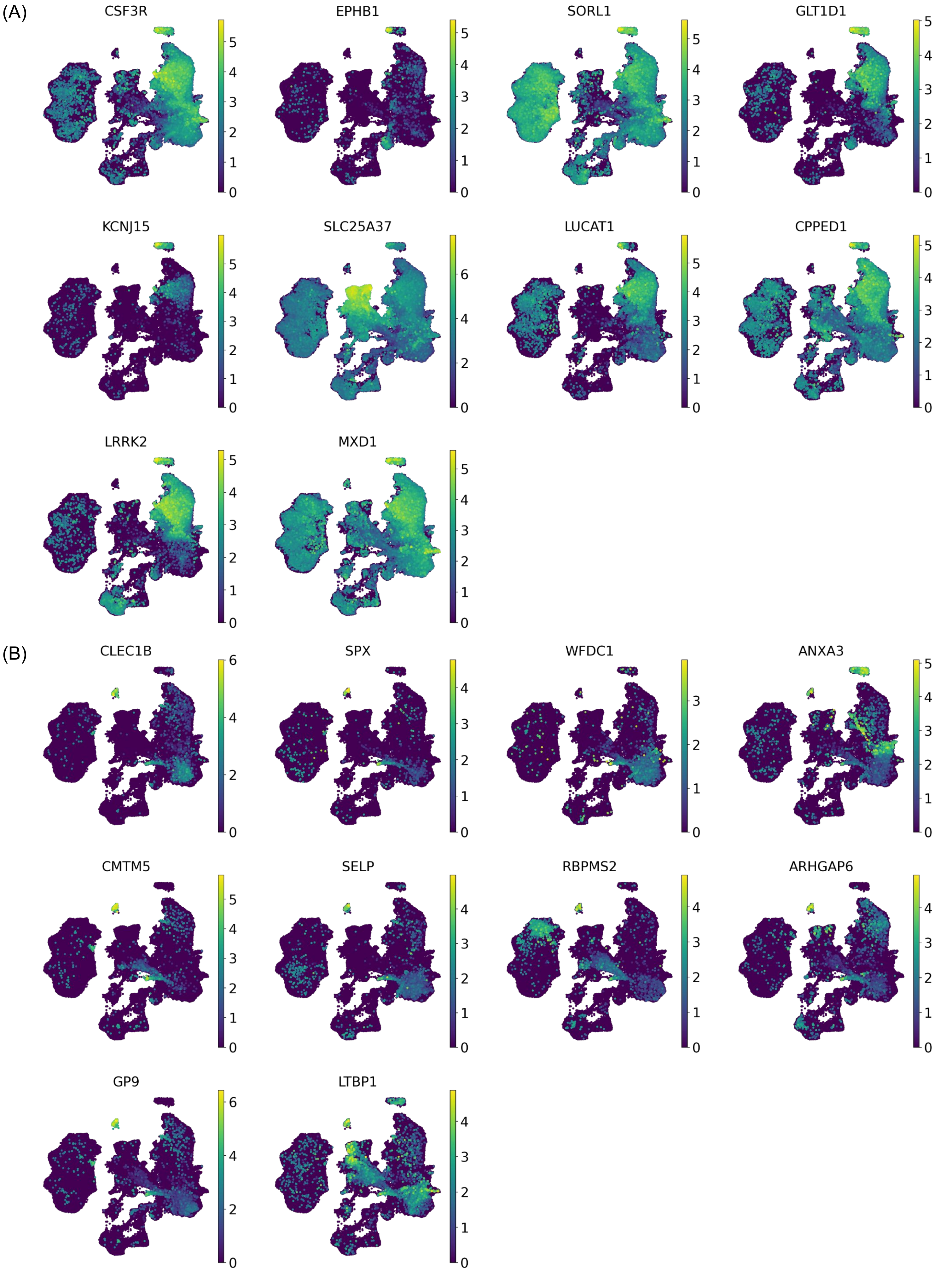


**Supplemental Figure 15. Expression of marker genes of cell types re-annotated by hand. (A)** Expression of neutrophil marker genes from Azimuth Human Adipose marker list[31]. **(B)** Expression of megakaryocyte progenitor marker genes obtained from Azimuth Human BM marker list[31].
