## Supplemental Data for "Distinct Stem Cell Identities Converge into Shared Erythroid Stress in ERCC6L2 Disease and Shwachman-Diamond Syndrome"

### Supplemental Methods

#### Sample collection

Patient information, including *TP53* mutation status, was collected from the Finnish Hematological Registry and Biobank – FHRB, and clinical repositories. Patients had a confirmed diagnosis of ERCC6L2 disease (ED) or SDS established through clinical genetic testing. The study was conducted in accordance with the Declaration of Helsinki. The study has been approved by Helsinki University Central Hospital ethics review committee (239/13/03/00/2010, 303/13/03/01/2011, 206/13/03/03/2016, amendment 2023, and HRHULAB2). All samples from living individuals are derived after written informed consent. ED and SDS patient samples are summarized in Supplemental Table 1 and detailed in Supplemental Table 2; AML patient samples are summarized in Supplemental Table 3.

#### Sample processing

##### ERCC6L2 disease and SDS

###### Fibroblasts and blood samples

Patient skin biopsies were obtained in conjunction with diagnostic samples. Fibroblast cultures were started from skin biopsies and cells were cultured with DMEM (Lonza, Basel, Switzerland) supplemented with 20% FBS (Gibco, Waltham, MA, USA) in the beginning, and 10% FBS once cell lines were established, as well as 1 % penicillin/streptomycin, and GlutaMAX (Gibco, Waltham, MA, USA), in 37°C, 80% humidity, 5% $CO_2$. Cells were passaged by trypsinization with 0.05% Trypsin-EDTA using standard methods.

Fibroblast cultures exposed to high (25 mM) or low (1 mM) glucose before RNA extraction to investigate the impact of glucose stress, as literature suggests slower growth rate of fibroblasts from patients with germline hematologic disorders[1], which we also observed (data not shown). Cells were collected by pelleting and lysed in Qiazol (Qiagen, Hilden, Germany). Samples were stored in -80°C until further processing.

Peripheral blood samples from patients and healthy controls were obtained in EDTA tubes and extracted with Nucleospin RNA Blood (Macherey-Nagel, Düren, Germany) according to the manufacturer's protocol. Total RNA was extracted from fibroblasts with miRNeasy Mini kit (Qiagen, Hilden, Germany) according to manufacturer's protocol.

RNA concentration and purity was measured using Nanodrop (Thermo Fisher Scientific, Waltham, MA, USA). RNA integrity was measured with Tapestation (Agilent, Santa Clara, CA, USA). Only samples with RIN>5 were included in this study.

The blood and fibroblast control samples were genotyped for the Finnish founder mutation at *ERCC6L2* (NM_020207.7):c.1424del (p.Ile475ThrfsTer36)[2–5] by Sanger sequencing from DNA extracted from whole blood (Nucleospin blood, Macherey-Nagel, Düren, Germany). All control samples were from donors of Finnish ethnicity, and confirmed as wild-type.

#### Bone marrow samples

1mL of bone marrow (BM) aspirate collected in EDTA-coated tube was incubated for 5 min at RT with 10mL of ACK lysis buffer (Thermo Fisher Scientific, A1049201) and centrifuged RT 5 min at 300 *x g* to remove the red blood cells. The ACK treatment was repeated until the cell pellet had no red residue. The cell pellet was resuspended in cold PBS (Gibco, 10010015) and centrifuged for 5 min at +4°C, 300 *x g*. For freezing, the cell pellet was resuspended in 10% DMSO-FBS (Thermo Fisher Scientific, 11402691; Thermo Fisher Scientific, 10270106).

The viably frozen cells were thawed in +37°C water bath for 2 min, followed by a modified 10X Genomics General Sample Preparation Protocol (CellPrepGuide_RevC); cells were sequentially diluted as advised in 10X Genomics PBMC protocol (FreshFrozenHumanPBMCs_RevE) and suspended in 2% BSA-PBS (Biotop, P6154-100). The dilution was centrifuged for 5 min at RT, 300 *x g* and washed twice with 2% BSA-PBS. The cell suspension was strained using a Flowmi Tip Strainer 40μM (Thermo Fisher Scientific, 15342931). Small amount of the suspension was dyed with trypan blue (Thermo Fisher Scientific, T10282) and the cells were counted on a chamber slide (Thermo Fisher Scientific, C10228) using Countess II Automated Cell Counter (Thermo Fisher Scientific, AMQAX1000). The cell concentration was adjusted to 700-1200 cells/μL. 0.2U/μL of RNasin

Ribonuclease Inhibitor (Promega, N2511) was added in cell suspension and the samples were placed on ice for transport and further processing.

#### Identification of somatic *TP53* variants

The identification of somatic mutations and their VAF in ED and SDS patient samples was performed as part of clinical diagnostics at the center of care of the patients. In brief, the targeted enrichment of 45 genes (all protein coding exons from the genes *ASXL1, BCOR, CDKN2A, CEBPA, CREBBP, CUX1, DDX41, DNMT3A, EP300, ETV6, EZH2, GATA2* (including conserved sequence from intron 4), *JAK2, KDM6A, NF1, PHF6, RAD21, RUNX1, SETD2, STAG2, TET2, TP53,* and *ZRSR2*, as well as targeted regions of genes *BRAF, CALR, CBL, CSF3R, FLT3, GATA1, IDH1, IDH2, KIT, KRAS, MPL, NPM1, NRAS, PDGFRA, PTPN11, SETBP1, SF3B1, SMC1A, SMC3, SRSF2, U2AF1,* and *WT1*) that are recurrently mutated in myeloid malignancies was performed using the Ion Torrent system with a custom primer panel followed by sequencing using the Illumina MiSeq (v.3 chemistry). Targeted areas were sequenced on average in 8000x depth, to assure >98 % of targeted read frames are sequenced at least 500x.

#### AML

Cryopreserved AML BM samples were obtained through the Finnish Hematology Biobank. Samples were processed and BM mononuclear cells (BMMC) were isolated by Ficoll-Paque density gradient (GE Healthcare).

### Bulk RNA sequencing and preprocessing

3’RNA-sequencing was performed for blood and fibroblast RNA. NEBNext Ultra Directional II kit (New England Biolabs, Ontario, Canada) was used for library preparation after rRNA depletion. Then, 3’ end labeling method based on Drop-seq protocol [6] was applied and sequencing was performed with Nextseq 500/550 (Illumina, San Diego, CA, USA), high output, 75 cycles.

A light quality trimming was applied to the data with Trimmomatic software[7]. The filtered reads were processed using Drop-seq tools v.2.3.0 according to the Drop-seq[6] pipeline and adapted to 3’ RNA-seq. The read libraries were converted to sorted, unaligned BAM files using Picard toolkit v.2.4.1[7].

Reads were tagged with sample-specific barcodes and unique molecular identifiers (UMIs), and 5’ adapters and 3’ poly A tails were trimmed. Reads were converted to FASTQ files and aligned with STAR v.2.7.6a[8]. Alignment was done using data from GENCODE Release 41 human genome GRCh38.p13[9] and comprehensive gene annotation files with default STAR settings. Uniquely aligned reads were sorted and merged with previous unaligned tagged BAM files to regain barcodes and UMI's lost during the alignment step. Annotation tags were then added to aligned and barcode-tagged BAM files.

Systematic synthesis errors in sample barcode sequences were detected and corrected with Drop-seq[6] tools. Digital expression matrices were created by counting the total number of unique UMI sequences (UMI sequences differing only by a single base are merged) for each gene.

### Single-cell RNA sequencing and preprocessing

#### ERCC6L2 disease and SDS

We applied single-cell RNA sequencing (scRNA-seq, 3' 10X Genomics) and targeted genotyping by single-cell amplicon sequencing (scAmp-seq) following established protocols[10,11] on BM samples from ED and SDS patients. The majority of *TP53* variants that the ED and SDS patients carry are missense variants, which are located mostly in the p53 DNA-binding domain, similarly to other haematopoietic neoplasms (Supplemental Figure 1). We genotyped all *TP53* mutations reported in the patients' myeloid panel except for splice variant c.376-2A>G for patient E6 (Supplemental Figure 1), which cannot be genotyped from mature mRNA reads. The somatic mutation pattern resembles other tumor types, where most somatic *TP53* variants found are missense mutations in the DNA-binding domain[12], affecting the DNA-binding ability or conformation of p53[12,13]. According to the clinical myeloid panel, neither the ED nor the SDS patients carry other somatic mutations that are recurrently mutated in myeloid malignancies. We perform no enrichment of any cell type and include all hematopoietic cell types in our analysis, except red blood cells.

For the ED and SDS samples, the Chromium Single Cell 3' Gene Expression run and library preparations were done using the 10X Genomics Chromium Next GEM Single Cell 3' Gene Expression version 3.1 Dual Index chemistry. Libraries were sequenced on Illumina NovaSeq 6000 platform using read lengths: 28bp (Read 1), 10bp (i7 Index), 10bp (i5 Index) and 90bp (Read 2). The resulting files were processed using CellRanger 6.0.2 pipelines with default parameters to generate FASTQ files and count matrices. The Illumina bcl2fastq v2.2.0 was used to run the mkfastq pipeline and alignment was done using GRCh38.

#### AML

For AML patients scRNA-seq was performed using the 10X Genomics Chromium Single Cell 3' Gene Expression version 2 (samples AML20 and AML21) and version 3 (all other AML samples) reagent kit. The manufacturer's instructions were followed for generating gel beads in emulsion (GEM), cDNA amplification, and library preparation. The prepared libraries were sequenced on the Illumina NovaSeq 6000 system. The AML samples were processed using the CellRanger 6.0.1 pipeline, and the AML M6 sample[14] was reprocessed using the CellRanger 6.0.2 pipeline.

#### Healthy controls

As healthy controls (HC) we used scRNA-seq data from 63 samples from 8 healthy adult human donors[15] that were obtained from the hematopoietic immune cell atlas[16] created for the Human Cell Atlas (HCA) project[17]. We reprocessed the HCA files using the CellRanger 6.0.2 pipeline.

### scRNA-seq data analysis and cell type identification

QC on scRNA-seq data was performed by Scanpy v.1.9.1[18]. We removed cells with more than 20% of mitochondrial transcripts (mtRNAs) as it has been shown that this is a sufficient threshold for human BM[19] and removed cells with less than 200 genes expressed to remove barcodes originating from droplets with ambient/cell-free RNA only[20]. We did not perform further filtering until necessary in downstream data analysis following recommendations[21].

Data integration and cell type identification was performed using Single-cell variational inference (scVI)[22] and Single-cell ANnotation using Variational Inference (scANVI)[23] as they have been shown to be top-performing on different complex integration tasks[24–26], in combination with a hematological marker database[27]. Both, scVI and scANVI were run with scvi-tools v.0.17.4 and scVI was setup with *sample* as batch variable, and *individual*, *condition* (ED, SDS, AML, and HC), diagnosis (healthy, BM normal, BMF, MDS, MDS/AML, AML) and *protocol* (Ficol, ACK) as categorical covariates. The scVI model was created with two hidden layers used for encoder and decoder NNs, 128 nodes per hidden layer and a dimensionality of 30 of the latent space, and gene expression is modeled by negative binomial distributions, following settings used in a recent integration of a large and diverse set of samples and cells across health and disease[26]. Covariates are concatenated to expression in encoder, but not to output of hidden layers in encoder and decoder, layer normalization is used in both, encoder and decoder, and no batch normalization is used. The scVI model is trained with a maximum of 500 epochs, with an early stopping criteria using evidence lower bound (ELBO) validation as stopping monitor, patience of 10, minimum delta of 0.0, and checking validation every epoch. The validation loss is monitored and the learning rate reduced by a factor of 0.1 after 8 epochs without improvement.

scANVI defines cells with top cell type score as seeds, where the cell type score of a cell is the sum of the normalized, logarithmized and scaled (unit variance and zero mean) expression values of that cell type's marker genes[23]. We used a hematological marker database [27] as cell type marker genes and approximately 0.1% of all cells (500 cells per cell type) as seeds. We initialized the scANVI model with weights from the pretrained scVI model and trained it with a maximum of 200 epochs, with an early stopping criteria using classification loss train as stopping monitor, training size of 1.0, patience of 10, minimum delta of 0.001, and checking validation every epoch. Finally, based on latent embedding obtained from scANVI we computed a neighborhood graph and generated a two-dimensional Uniform Manifold Approximation and Projection (UMAP)[28] for visualization using Scanpy v.1.9.1[18]. The identified 32 different cell types include all haematopoietic cell lineages (Figure 1C), which is comparable to other recent single-cell studies of the BM[29,30]. Quality control (QC) of the data showed expected differences between the conditions, such as an accumulation of hematopoietic stem cells (HSCs) and early progenitor cells in AML compared to the other conditions (Supplemental Figure 7).

We noted that some myelo- and monocytes located in the UMAP separately from most myelo- and monocytes, which consisted of cells from ED and SDS patients only, whereas the other myelo- and monocytes consisted of cells from all conditions. To re-evaluate, which cell type those represent, we clustered them using Leiden[31] (resolution 0.08) and performed a Wilcoxon rank-sum test with Scanpy's rank_genes_groups function (separately located myelo- and monocytes against all other cells, except the other myelo- and monocytes) with default parameters. We then performed pathway enrichment using enrichR with the identified upregulated DEGs ($p_{adj}$<=0.05 and log2FC>=1) as input. The most enriched cell type identified were neutrophils from the Azimuth's adipose cell type list, and neutrophil marker genes – obtained from the Azimuth Human Adipose marker list[32] – were highly expressed in the separately located cells (Supplemental Figure 15A, Supplemental Tables 27-28). Neither Azimuth's BM nor Triana's marker list include neutrophils as cell type, as they are discarded when using Ficoll (used for HD and AML samples) to extract mononuclear cells, whereas neutrophils are included when using ACK (used for ED and SDS samples). Hence, we annotated these cells as neutrophils.

Further, two distinct populations were annotated as megakaryocyte progenitors. Clustering them using Leiden[31] (resolution 0.08) and performing Wilcoxon rank-sum test and enrichR (with settings as with the myelo- and monocytes), the top two enriched cell types identified were platelets and megakaryocyte progenitors for both populations, and megakaryocyte progenitor marker genes – obtained from Azimuth Human BM marker list[32] – were expressed in both (Supplemental Figure 15B, Supplemental Table 27-28). Hence, we annotated these cells as Megakaryocyte progenitor 1 (MkP1) and Megakaryocyte progenitor 2 (MkP2).

### Single-cell amplicon sequencing

To identify the *TP53* mutation status of single cells, we used custom primers for 3' 10x Genomics to amplify the *TP53* variant positions (Supplemental Table 4), performed QC, and identified the *TP53* mutation status for reads, mRNA transcripts, and finally cells(Supplemental Figure 3A).

More specifically, we followed a targeted genotyping protocol outlined by Van Egeren et al.[11] with slight modifications to enrich *TP53* gene transcripts from 10x Genomics Chromium Single Cell 3' Gene Expression amplified cDNA. Locus-specific single cell amplicon (scAmp-seq) libraries were generated using a double-nested PCR approach[11] resulting in four PCR rounds. Locus-specific reverse primers preserving the mutation site together with generic forward primers preserving the single-cell barcoding structure were designed (Supplemental Table 4). Reaction volumes were 20 µl in all except the 4th PCR where 25 µl was used. All PCR reactions were purified using 0.8x SPRIselect (Beckman Coulter #B23318) after each amplification was completed.

In Step 1, a 20 µl PCR reaction contained 10 µl 2x KAPA Hifi PCR Master Mix (Roche #KK2602), 1.2 µl forward primer (SI-PCR, Illumina P5 sequence and partial Read1) at 10 uM, 0.6 µl reverse primer (scAmp_TP53_ex1_1 or scAmp_TP53_ex2-3_1) at 10 µM, 3 ng of full length 10x cDNA and remaining volume was added up with nuclease-free water. The reactions were amplified using the following protocol: initial denaturation at 95°C for 180 s, 15 cycles of denaturing at 98°C for 20 s, annealing at 67°C for 30 s, extension at 72°C for 120 s, and a final extension at 72°C for 180 s.

In Step 2, a 20 µl PCR reaction contained 10 µl 2x KAPA Hifi PCR Master Mix (Roche #KK2602), 0.6 µl forward primer (Partial P5) at 20 µM, 0.6 µl reverse primer (scAmp_TP53_ex1_2 or scAmp_TP53_ex4_1) at 10 µM, 2 µl of purified product from Step 1 and 6.8ul of nuclease-free water. The reactions were amplified using the following protocol:

initial denaturation at 95°C for 180 s, 15 cycles of denaturing at 98°C for 20 s, annealing at 67°C for 30 s, extension at 72°C for 120 s, and a final extension at 72°C for 180 s.

In Step 3, a 20 µl PCR reaction contained 10 µl 2x KAPA Hifi PCR Master Mix (Roche #KK2602), 0.6 µl forward primer (Partial P5) at 20 µM, 0.6 µl reverse primer (amplicon specific, scAmp1_TP53, scAmp3_TP53, scAmp4, _TP53, scAmp5_TP53) at 10 µM, 1.6 µl of purified product from Step 2 and 7.2 µl of nuclease-free water. The reactions were amplified using the following protocol: initial denaturation at 95°C for 180 s, 10 cycles of denaturing at 98°C for 20 s, annealing at 67°C for 30 s, extension at 72°C for 120 s, and a final extension at 72°C for 180 s.

In Step 4, a 25 µl PCR reaction contained 12,5 µl 2x KAPA Hifi PCR Master Mix (Roche #KK2602), 0.5 µl forward primer (SI-PCR) at 10 µM, 2.5 µl reverse primer (Single Index Kit T Set A, #PN-1000213, 10x Genomics), 2 µl of purified product from Step 3 and 7.5 µl of nuclease-free water. The reactions were amplified using the following protocol: initial denaturation at 95°C for 180 s, 10 cycles of denaturing at 98°C for 20 s, annealing at 54°C for 30 s, extension at 72°C for 120 s, and a final extension at 72°C for 180 s.

### scAmp-seq preprocessing, data analysis and identification of *TP53* mutation status

scAmp-seq libraries were sequenced on the NovaSeq platform. The resulting files were processed using CellRanger 6.0.2 with default parameters to generate FASTQ files and count matrices. We analyzed the FASTQ files of the scAmp-seq libraries to identify individual cells as either *TP53* wild-type (WT) or *TP53* mutant (MUT):. First, we performed quality control (QC), refined from QC of previously published pipelines[10,11], then used genotype likelihoods[33] to define first reads, then mRNA transcripts and finally cells as WT or MUT (Supplemental Figure 3A). Next, we describe each step in more detail.

First, our scAmp-seq data analysis pipeline performed quality control (QC): We performed barcode QC by keeping only those barcodes that occur on the 3M-february-2018.txt.gz barcode whitelist provided by 10x Genomics' CellRanger v.6.0.2. For sequencing QC reads in the FASTQ files were discarded if the average Illumina base quality value was less than 30 as in Van Egeren et al.[11]. We do not correct barcodes or UMIs as a recent benchmark shows that only few additional reads (on average, 0.8% of the reads with Hamming distance=1, and additional 0.0038% with Hamming distance=2) are rescued[34]. For primer QC reads were kept only if the primer sequence read includes known primer sequence exactly or with maximum one base pair mismatch similar to Nam et al.[10]. Next, we performed read QC. In case of single nucleotide variant (SNV) substitutions, we retained reads which have WT or MUT base at variant position and which neighborhood (+/-10bp of variant position) matches *TP53* CDS reference exactly or with maximum one base pair mismatch similar as in Van Egeren et al.[11]. In case of indels (insertion or deletion variants), we retain reads which match either the expected mutated *TP53* sequence or *TP53* CDS reference exactly. As indels are harder to identify we do not accept a base pair mismatch for these. By doing so, we defined each read as WT or MUT. For mRNA transcript QC we kept only those mRNA transcripts (barcode-UMI pairs) for which there were at least two reads.

Second, we defined each mRNA transcript (barcode-UMI-pair) as WT or MUT. To identify *TP53* mutation statuses of mRNA transcripts, we used genotype likelihoods[33]. Therefore, we created a FASTQ file for each mRNA transcript, consisting of all reads associated with that mRNA transcript. We aligned the reads of each mRNA transcript to GRCh38 (creating a BAM file) using STAR v.2.7.9[8]. We then created an index BAI file using SAMtools v.1.13 [33], a variant call format (VCF) file using BCFtools v.1.9[33], and performed variant calling using BCFtools v.1.9[33] with parameter --ploidy 1 to assess the haploid variant status as a mRNA transcript represents a single mRNA molecule.

Third, we define cells (barcodes) as WT or MUT. We defined cells as WT w.r.t. to a specific variant if <10% of their mRNA transcripts (barcode-UMI -pairs) were WT, and as MUT if >30% of their mRNA transcripts were MUT. For those cases where the *TP53* variant position was covered by reads produced with two alternative primers (scAmp5_I and scAmp5_II, or scAmp4 and scAmp5_I) we combined the data produced with the two primers as followings: *TP53* mutation status for cells with identical *TP53* mutation status for both primers and cells with *TP53* mutation status for only one primer were kept, whereas *TP53* mutation status for cells with conflicting *TP53* mutation statuses (that is, *TP53* WT for one primer, but *TP53* MUT for the other primer) were removed from downstream analysis.

Finally, in contrast to the previously published pipelines which identify mutation statuses of single cells for one variant per gene[10,11,30], our study also includes cases where patients carry up to four *TP53* variants. Hence, cells need to be defined as WT or MUT w.r.t. multiple variants. The steps described above were performed for each variant separately. Afterwards, the mutation status of the different variants were combined as follows: a cell was defined as *TP53* WT only if for each *TP53* variant of the sample the mutation status of the cell was WT whereas the cell was defined as MUT if the mutation status of at least one *TP53* variant was MUT. For cells, where the mutation status of one variant is wild-type, but the mutation status information is missing for another variant locus, we cannot define the cell's overall *TP53* mutation status. Consequently, the fraction of cells with an overall *TP53* mutation status per sample is smaller than the sum of the fractions over all that patient's *TP53* variants. Applying this approach, we identify the *TP53* mutation status in 0.8–59.5% of cells per sample, mutation site and primer, in 1.5–61.4% of cells per sample and mutation site (Supplemental Figure 3B), and in 2.1–44.6% of cells per sample (Supplemental Figure 3C).

We classified all cells from patients E2, SDS1 and SDS2 as WT since no somatic *TP53* variants were detected in the patients' clinical myeloid mutation panel. We mapped the TP53-mutated and wild-type status of cells onto the scRNA-seq data using the shared single-cell barcodes by adding the *TP53* mutation status information as observations to the anndata object obtained from the scRNA-seq data. Neither the primer choice nor the viability of the cells did seem to affect the number of cells for which the mutation status was identified (Supplemental Figure 3D). Further, running our pipeline on a known WT position showed that it identifies few false positives at the read level, and almost no false positives at the mRNA transcript and cell levels (Supplemental Figure 3E).

Previous approaches have identified the mutation status for highly expressed genes such as *CALR* in over 90% of cells[10,30]. However, identifying the mutation status of low expressed genes such as *TP53* has been shown to be challenging[30,35] , identifying them in less than 5%–75% of cells for *TP53*[30,36] and in 5%–15% of cells for *JAK2*[11]. Of note, these previous publications focus on myeloproliferative neoplasms (MPN)[10], polycythemia vera and essential thrombocythemia[11], where *JAK2* variant leads to an activated JAK2 signaling, or on AML[30] and secondary AML (sAML)[36], where gene expressing elevated compared to non leukemic cells. Recent pipelines analyze variants in several genes per patient and reconstruct clonal hierarchies[30,35,36]. However, these clonal hierarchy reconstruction methods are restricted to variants with coverage in at least 20% of cells[35] or in at least 60% of cells for one variant[30], making these methods not applicable for our BMF samples with small *TP53* clones.

Taken together, we identified the *TP53* mutation status at variant level comparable to previous studies when taking into account that we assess the mutation status of a low expressed gene mostly in BMF and MDS, and combine mutation status of different variants to a *TP53* mutation status at cell level.

### Lollipop plots

Lollipop plots were generated with ProteinPaint[37,38]

### Examination of *ERCC6L2* raw reads

To determine whether the mRNA transcripts of *ERCC6L2* carrying the deletion variant can be observed in our scRNA-seq data we examined the aligned sequences (BAM files) of the raw reads provided by 10x Genomics using BasePlayer v.1.0[39] and checked the *ERCC6L2* c.1424delT variant position. Although 3' 10x Genomics is not a full-length transcriptome method, the obtained scRNA-seq data contains reads along whole transcripts[40,41]. The main transcript of wild-type *ERCC6L2* (ENST00000653738.2) has 19 exons. In our data, the *ERCC6L2* germline variant c.1424delT (p.Ile475ThrfsTer36, rs768081343), a single nucleotide deletion in exon 9, is present in the ERCC6L2 patients' scRNA-seq reads aligned to the reference of the *ERCC6L2* gene (Supplemental Figure 6B).

### Differential expression analysis for bulk RNA-seq

All differential expression analyses were performed using DESeq2 v.1.40.1[42] using default parameters (Wald-test, α=0.05). No difference between different media was found in the fibroblasts within a disease group, thus further analysis was carried for pooled samples within a disease group in fibroblasts. Blood samples were controlled for whether patients carried at least one *TP53* clone (yes/no) and sex. ED and SDS fibroblasts were controlled for media, sex, batch and time point.

### Differential expression analysis for scRNA-seq

To identify differentially expressed genes we first normalized the data with a scale factor of 10,000, thereby excluded very highly expressed genes (more counts than 1% of the total counts in at least one cell) from the computation of the size factor for each cell as these can strongly influence the resulting normalized values for all other genes[21,43]. To do so, we used Scanpy's normalization function (with target_sum=1e4, exclude_highly_expressed=True, max_fraction=0.01). We then examined genes that are expressed in at least 10% of cells in a cell type and identified DE genes for each cell type separately by MAST v.1.24.1[44] using its two-part generalized linear model (zlm function with method='bayesglm') and a likelihood-ratio test to compare the condition we are interested to a reference condition, thereby setting cellular detection rate (CDR)[44] and age at sampling as continuous covariates and sex as factor.

### Expression of *ERCC6L2*, *SBDS*, and *TP53*

DESeq2 only identifies genes as differentially expressed with counts > 10, thus we used Python's SciPy v.1.10.1 one-way ANOVA and statsmodels v.0.13.5 Tukey's test to compare normalized expression of *ERCC6L2*, *SBDS*, and *TP53* in blood, BM and BM (Supplemental Figure 6A).

### Pathway analysis

To identify enriched pathways we performed pathway analysis using enrichR[45] using Reactome_Pathways_2024[46] gene sets. DE genes with $p_{adj}$ < 0.05 and absolute log-fold change |log2FC| > 0.5 were considered significant, and were included in pathway analysis of DE genes.

### Pseudotime analysis

To compare differences in gene expression dynamics along differentiation between conditions we calculated the diffusion pseudotime[47] and calculated expression dissimilarities along the pseudotime trajectory. We first normalized the data with a scale factor of 10,000 and then log transformed it using Scanpy v.1.9.1[18]. To select root cells for the pseudotime calculation we performed cell scoring implemented in Scanpy v.1.9.1[18] based on HSC & MPP marker genes from a hematological marker database[27]. The pseudotime was calculated with CellOracle[48], which uses Scanpy's implementation of the Diffusion Pseudotime (DPT) from[47] for each lineage separately.

To compare the pseudotime trajectories of cells in different conditions we compared the genewise expression along the pseudotime trajectory. We binned the gene expression with pseudotime trajectory alignment tools from Genes2Genes (G2G)[49]. The gene expression was normalized by the average gene expression of the healthy controls. To quantify the difference in gene expression between two groups we calculated the absolute difference in gene expression in each bin. We took the mean of these differences and denoted it as the expression dissimilarity between two groups. To obtain confidence intervals and mean for the expression dissimilarity, we performed bootstrapping: We resampled the cells 100 times from the original dataset, rerun the alignments, and collected the expression dissimilarity for each gene for each run. In the end the 95% confidence interval and mean are calculated from all these runs.

### Erythroid, cell cycle and cell death scores

The scores were calculated by Scanpy's score_genes function with previous published gene lists: an erythroid erythroid-curated[36], a cell cycle[36], pro- and anti-apoptotic[50] and ferroptosis[51] gene list, of which the latter is also found in the ferroptosis pathway in KEGG[52]. Genes included in these gene lists can be found in Supplemental Figures 11-12.

### Cell cycle analysis

To identify cell cycle phases of individual cells we first normalized (using a scale factor of 10,000), log transformed and scaled the data using Scanpy v.1.9.1[18] and then used cell cycle scoring[53] implemented in Scanpy v.1.9.1[18] with previously defined cell cycle genes[54]. To assess whether cells in one condition are preferentially more often in a cell cycle phase than in another condition, we performed bootstrapping with 10,000 resamples, and calculated confidence intervals as well as the fraction of resamplings in which the fraction of cells of one condition in a cell cycle phase is higher than the fraction of cells of another condition in the same cell cycle phase.

### Transcription factor and gene regulatory network analysis

We regressed out the *TP53* mutation status, cell cycle scores and potential confounders sex and age using Scanpy v.1.9.1[18]. Differentiation trajectories were inferred using PAGA[55] for each condition separately. To obtain CIs for the HSC & MPP proportions and *p*-values for their comparisons we performed bootstrapping with 10,000 resamples. HSC & MPP were clustered using Leiden[31] (resolution=0.5). Transcription factors (TFs) were obtained from a recent publication[56] in order to calculate the Pearson correlation between differentially expressed TFs and non-TF DEGs using the Python scipy v.1.10.1 package. We built the gene regulatory network (GRN) using CellOracle v.0.12.1[48] and plotted a subnetwork of the GRN containing the differentially expressed TFs using python-igraph v.0.10.4[57].

### Single cell copy number analysis with inferCNV

We used inferCNVpy v.0.5.0, a Python reimplementation of inferCNV[58], to identify copy number variants (CNVs) in the scRNA-seq data for the ED MDS/AML (E10) and SDS MDS (SDS5b) samples. CNVs were detected per cell type with a window size of 250 and using an average of the expression of cells from 20 healthy controls as reference. Dimension reduction was performed with PCA and cells were clustered based on their CNV patterns using Leiden[31] (resolution 0.35), both implemented in the inferCNVpy package.

### Gradient boosting on BMF stable/acceleration status

We investigated whether disease acceleration in ED blood or BM prior to malignant transformation could be detected. We applied gradient boosting[59] on our transcriptomics data. We investigated if genes with most variable expression would reveal differences between BMF samples with clinically, genetically, and morphologically stable disease (BMF-stable (Supplemental Table 2); 12 blood samples from five patients, six BM samples from two patients), and BMF samples with signs of disease acceleration (BMF-acceleration (Supplemental Table 2); six blood samples from four patients and three BM samples from three patients). Acceleration was defined as increasing erythropoiesis in adjacent BM biopsy and increase in the VAF or number of *TP53* clones[3]. We used only samples from adult female patients for the prediction, because our data comprises a limited number of male and pediatric patients.

We removed mitochondrial genes (mt) genes, hemoglobin genes, and genes in the X and Y chromosomes from the expression dataset. The mt-genes were removed to account for the effects of their high expression levels, and potential mitochondrial heteroplasmy. The hemoglobin genes were removed as they can similarly have very high expression counts and thus cause biases in downstream analyses. The genes from X and Y chromosomes were removed due to their sex specific expression levels. Additionally, we removed alternative transcripts to avoid duplicate genes by removing all genes with a '.' in the name. In total, expression data for 33,760 and 21,419 genes was accounted for in blood and BM, respectively. For the scRNA data we summed sample-wise gene counts on erythropoietic cells alone and summed the sample-wise gene counts across HSC & MPP, EMP, EEP, and LEP. Normalization for the gene expression levels was conducted as follows: first we incremented all values by one. Second, the expression level of a gene was divided by the total sum of gene expression levels (read counts) per sample. Finally, we controlled for the batch with ordinary least squares regression. We created an individual model for each gene where the predictors were defined as batch and the predicted value was defined as the gene expression level, and obtained adjusted expression values by subtracting the predicted values from original values. For the following analyses, we chose a fraction of the most variable genes, ranging from 1% to 10% with 1% increments.

We used the clustermap function in the Python seaborn package v.0.12.2[60] to cluster the normalized gene expression data from the samples in order to investigate whether there were differences between the sample groups (Supplemental Figure 5). We then constructed models to predict if a sample belonged to the BMF stable or BMF acceleration group based on the normalized gene expression data using Python XGBoost v1.7.6[59] package (Supplemental Figure 5). We also shuffled the group labels to examine performance on data with no correlation between the labels and expression data (Supplemental Figure 5). Due to the small amount of data, we performed a leave-one-out cross validation with seven folds for bulk RNA analysis and five folds for scRNA-analysis. In each fold, an XGBoost model was trained on six patients' bulk RNA-analysis and on four patients' scRNA-analysis data and tested on the single remaining patient's data. We used the default parameters of the XGBoost package, except for the importance type which was set as "gain". Model accuracy was evaluated by calculating the mean of the individual fold accuracies from the leave-one-out cross validation.

### Statistical analysis

Fisher's exact t-tests were performed using R v.4.1.3, and q-values (adjusted p-values) were computed using the Benjamin and Hochberg method in either R or Python.

55. Wolf FA, Hamey FK, Plass M, et al. PAGA: graph abstraction reconciles clustering with trajectory inference through a topology preserving map of single cells. *Genome Biol*.

2019;20(1):59. doi:10.1186/s13059-019-1663-x

56. Lambert SA, Jolma A, Campitelli LF, et al. The human transcription factors. *Cell*. 2018;172(4):650-665. doi:10.1016/j.cell.2018.01.029

57. Csardi G, Nepusz T. The igraph software package for complex network research. *InterJournal, Complex Systems*. 2006;1695.

58. Patel AP, Tirosh I, Trombetta JJ, et al. Single-cell RNA-seq highlights intratumoral heterogeneity in primary glioblastoma. *Science*. 2014;344(6190):1396-1401. doi:10.1126/science.1254257

59. Chen T, Guestrin C. XGBoost: A Scalable Tree Boosting System. In: *Proceedings of the 22nd ACM SIGKDD International Conference on Knowledge Discovery and Data Mining*. ACM; 2016. doi:10.1145/2939672.2939785

60. Waskom M. seaborn: statistical data visualization. *J Open Source Softw*. 2021;6(60):3021. doi:10.21105/joss.03021
